# Motor cortex pain compensation is modifiable: allostatic load degrades its analgesic efficiency and rehabilitation restores it

**DOI:** 10.64898/2026.09.21.26363555

**Authors:** Felipe Fregni, Marta Imamura, Lucas Camargo, Kevin Pacheco-Barrios, Anna Carolyna Lepesteur Gianlorenço, Linamara Battistella, Wolnei Caumo

## Abstract

The primary motor cortex is a critical hub for pain control, yet its few identified regulatory mechanisms — intracortical inhibition and oscillatory markers — predict symptoms inconsistently. The prevailing view holds that pain results from reduced inhibition, a fixed deficit, rather than from resistance to inhibition that rehabilitation can reverse. Here we show that motor cortex pain compensation is instead a modifiable state whose analgesic efficiency depends on allostatic load. In a fibromyalgia trial and a knee osteoarthritis cohort assessed with parallel EEG and transcranial magnetic stimulation, a signature combining central theta asymmetry and intracortical inhibition was unrelated or inversely related to symptom burden under low allostatic load but positively related to it under high load in fibromyalgia, and its protective association was attenuated under high load in osteoarthritis — the same signal carrying different clinical meaning. A burden-adjusted index modelled on the homeostatic model assessment of insulin resistance was elevated under high load, predicted improvement after rehabilitation beyond baseline severity, and fell in responders but not in non-responders; in both cohorts the neurophysiological component itself recalibrated with response. Motor cortex compensation is thus degraded by allostatic load and restored by rehabilitation.

## INTRODUCTION

The primary motor cortex (M1) is increasingly recognised as an important node in pain modulation. Beyond its canonical role in motor control, M1 participates in endogenous pain regulation through descending projections involving the periaqueductal grey and rostral ventromedial medulla, thalamo-cortical loops, extrapyramidal pathways, and cortico-limbic circuits. Consistent with this broader regulatory role, both direct stimulation and behavioural engagement of M1 can produce clinically meaningful analgesia across chronic pain conditions.(1) Yet relatively few M1-related regulatory mechanisms have been characterised in patients — principally intracortical inhibition, cortical oscillatory activity, and their relationship to conditioned pain modulation — and their associations with pain severity and clinical outcomes have been inconsistent across individuals and studies.(2–4) These inconsistencies have largely been attributed to methodological heterogeneity, and an umbrella review of 49 evidence syntheses found that no cortical or peripheral measure had achieved clinical validation in chronic pain.(5) An alternative explanation is mechanistic rather than methodological: the same cortical signal may not carry the same meaning in every patient. Two observations point in this direction. Composite cortical signatures combining peak alpha frequency and corticomotor excitability predict experimental pain sensitivity but not clinical response to treatment,(6) suggesting that they capture engagement of a process rather than its effectiveness; and depression has been reported, at trend level, to modify the association between peak alpha frequency and pain,(7) suggesting that comorbidity alters the clinical meaning of a cortical signal rather than being only a confounder. Yet comorbidity is still treated predominantly as a source of confounding, and reduced cortical inhibition is still interpreted as a deficit, rather than as a compensatory response whose effectiveness depends on the physiological context in which it operates.

We hypothesised that the clinical meaning of a neural compensatory response depends on the individual’s available neurocompensatory reserve — the capacity of the neuroendocrine, immune and autonomic systems on which cortical regulation depends to translate compensatory engagement into effective pain control, a capacity that is progressively eroded by cumulative allostatic load.. A conceptual precedent exists in neurodegenerative disease, where compensatory neural activity can moderate the relationship between pathology and function.(8) Importantly, compensatory processes may intensify as pathology accumulates but subsequently decline once compensatory reserve is exhausted, such that similar levels of a compensatory parameter may represent different physiological states at different stages of disease.(9) In chronic pain, allostatic load — the cumulative physiological burden arising from repeated or insufficiently terminated activation of neural, endocrine, metabolic, and immune regulatory systems — provides a plausible mechanism through which such reserve may be progressively constrained.(10, 11) An analogous principle is well established in metabolic physiology: elevated insulin in the presence of preserved glycaemic control reflects effective compensation, whereas elevated insulin accompanied by hyperglycaemia indicates reduced regulatory efficiency or insulin resistance.(12) By analogy, increased cortical compensatory engagement may indicate either successful pain regulation or increasing compensatory saturation, depending on the systemic context in which that engagement occurs. Failure to account for this context dependence could therefore contribute to apparently inconsistent associations between neurophysiological markers and clinical outcomes and may limit the translational utility of cortical biomarkers in chronic pain.

Within this framework, the capacity of the motor cortex, an important hub for pain modulation, to regulate pain is conceptualised not as a fixed property, but as a dynamic resource whose efficiency depends partly on cumulative allostatic load across interacting physiological systems.(13) Depression, sleep disturbance, obesity, and perceived stress — each associated with altered pain regulation — converge on central pain-modulatory systems and may progressively constrain neurocompensatory efficiency.(14–17) We therefore propose that increasing allostatic load alters not necessarily the magnitude of compensatory engagement, but the relationship between compensatory engagement and its clinical effect. Under relatively low load, greater engagement may accompany effective pain regulation; under high load, comparable engagement may instead coexist with persistent symptoms, consistent with reduced compensatory efficiency. This framework provides a potential explanation for why similar neurophysiological signatures may have different, or even opposing, clinical associations across individuals. It further suggests that characterisation of neurocompensatory reserve may provide information that is not captured by symptom severity alone when developing biomarkers or identifying mechanisms relevant to treatment response.

To operationalise this framework, we developed a motor cortex-derived measure of cortical pain-compensatory resistance, the Cortical Pain-Compensatory Resistance Index (CPRI; Figure 1 and Table 1), conceptually modelled on the Homeostatic Model Assessment of Insulin Resistance (HOMA-IR; Table 2).(18, 19) M1 was selected because of its established position within the pain connectome and its involvement in descending inhibitory, thalamo-cortical, and cortico-limbic mechanisms implicated in analgesia.(1, 20) Central theta activity was selected because converging observations from our group suggest that slow-frequency theta activity may represent a compensatory neurophysiological response in chronic pain whose relationship with clinical status varies according to systemic burden.(21, 22) Left-hemisphere lateralisation was specified a priori based on evidence suggesting left-sided predominance during early nociceptive processing,(23) while recognising that the role of hemispheric lateralisation in chronic pain remains still under investigation and is examined in Methods.

**Figure 1.**
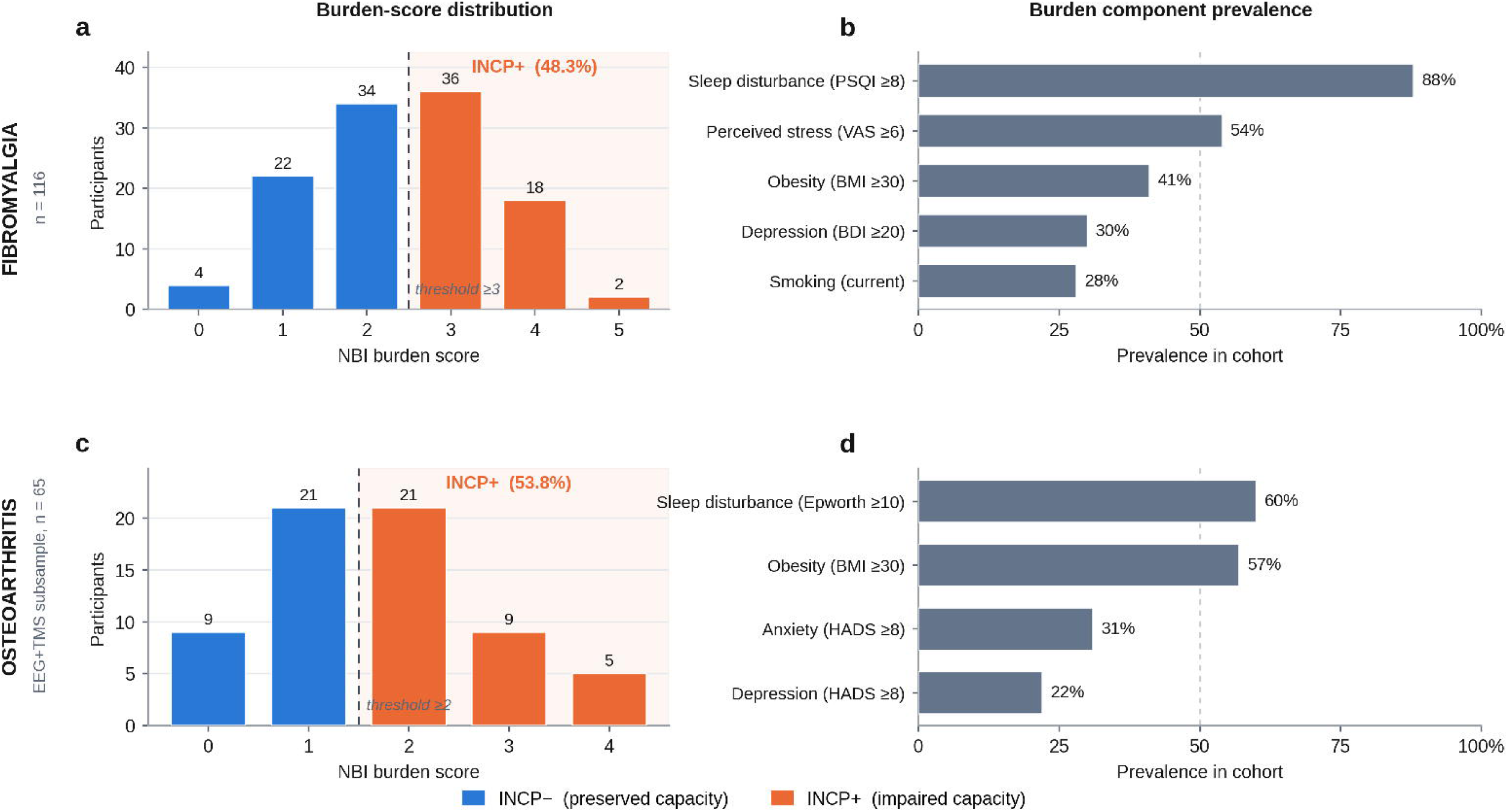
Conceptual model, empirical basis and index construction. a) Conceptual model. Cortical compensatory engagement (PCS) plotted against symptom burden under low (blue, INCP−) and high (orange, INCP+) allostatic load. Equivalent engagement carries opposite clinical association depending on load: under low load greater engagement accompanies lower symptom burden, whereas under high load comparable engagement coexists with persistent symptoms. Rehabilitation (green) restores the efficiency with which engagement translates into relief. b, c) Empirical basis in fibromyalgia. Associations of b) central theta asymmetry (left − right, z-scored; n = 104) and c) short-interval intracortical inhibition (SICI, z-scored; n = 113) with FIQR according to INCP status. Points represent participants, lines represent simple-slope regression estimates within each group, and shading indicates 95% confidence intervals; boxes report group slopes and the Predictor × INCP interaction from OLS models with HC3 robust standard errors. In b one INCP− participant (z = −6.5) lies outside the plotted range. d) Index construction. The Pain Compensatory System (PCS) index averages within-cohort z-scores of central EEG theta asymmetry and reverse-coded SICI (−SICI); higher PCS denotes greater left-lateralised theta activity and stronger inhibition. Symptom burden (FIQR in fibromyalgia; SF-36 Physical Functioning, reversed, in osteoarthritis) is z-scored within cohort. Both components are shifted to non-negativity and their product normalised to the cohort mean, giving the Cortical Pain-Compensatory Resistance Index (CPRI), with 1.0 denoting the cohort-average compensatory–symptom mismatch. Allostatic load, operationalised as the Neurocompensatory Burden Index (NBI) and dichotomised as INCP status, is the moderator and is not a term in the index. CPRI, Cortical Pain-Compensatory Resistance Index; FIQR, Fibromyalgia Impact Questionnaire-Revised; INCP, Impaired Neurocompensatory Capacity Phenotype; NBI, Neurocompensatory Burden Index; OLS, ordinary least squares; PCS, Pain Compensatory System; SICI, short-interval intracortical inhibition.

**Table 1.**
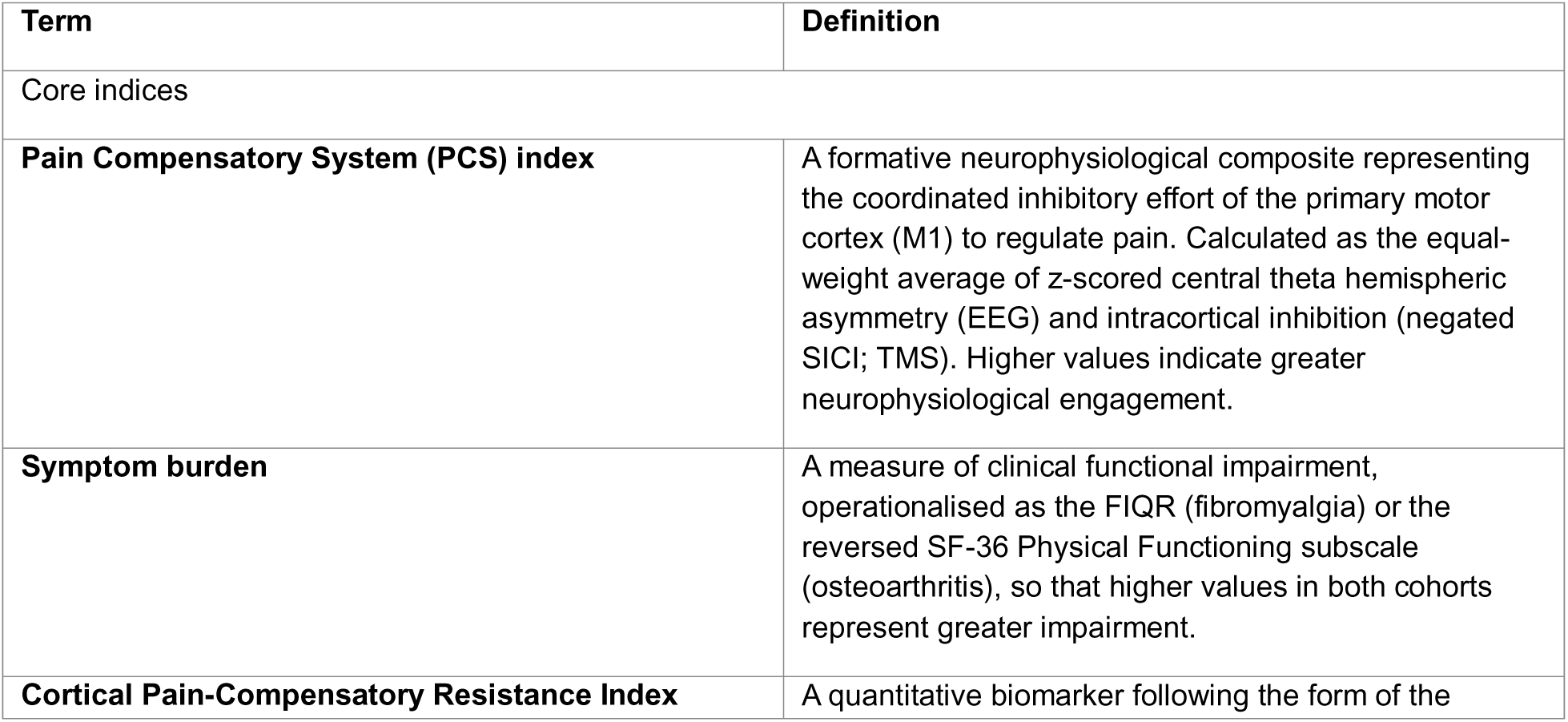
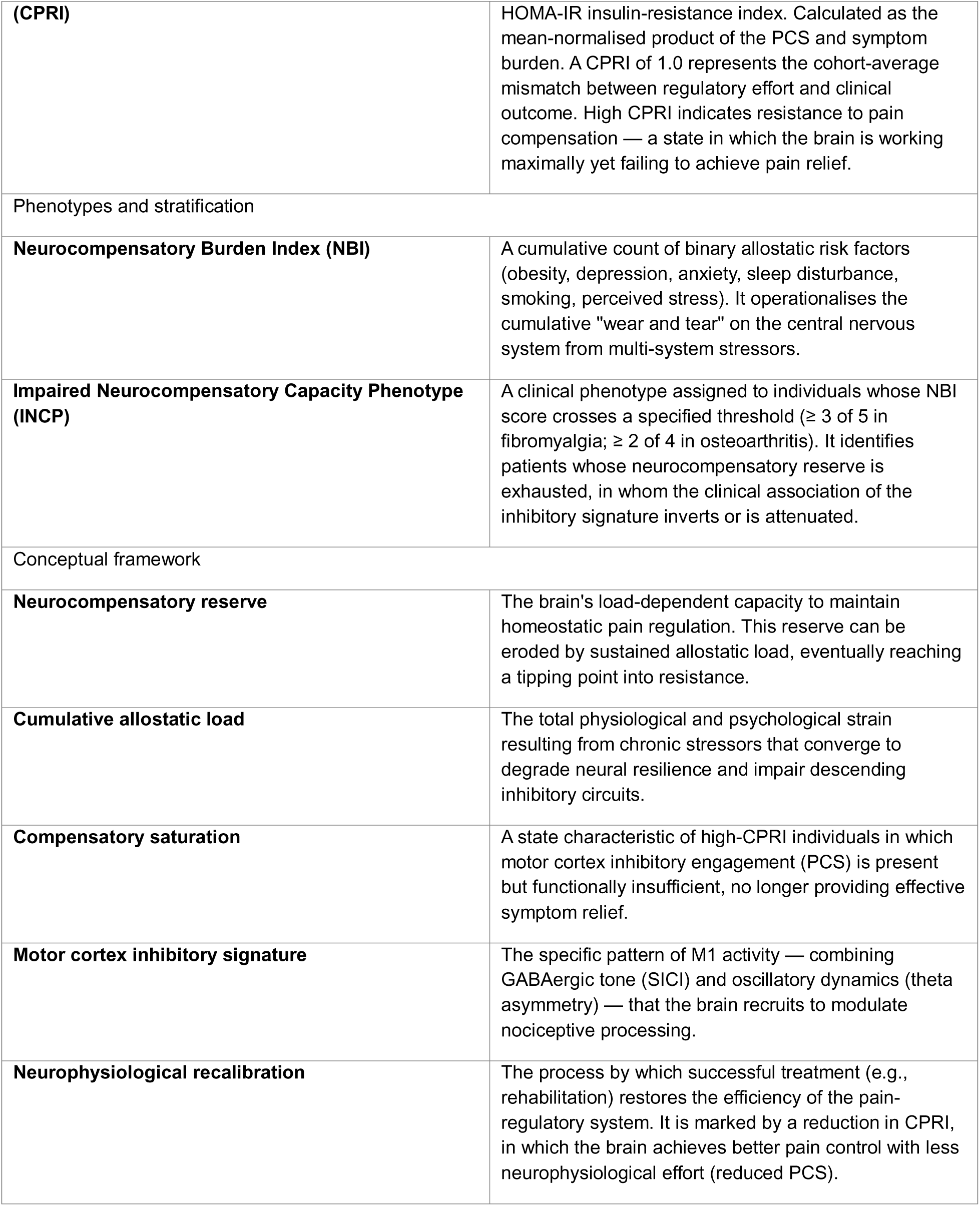
Glossary of the cortical pain-compensatory resistance framework.

**Table 2.** Comparison of the metabolic and cortical resistance frameworks.

| Feature | HOMA-IR (insulin resistance) | CPRI (cortical pain-compensatory) |
| --- | --- | --- |
|  |  | **resistance)** |
| **Core logic** | A high regulatory signal (insulin) indicates failure, not efficiency, in the presence of high output (glucose). | High regulatory effort (PCS) indicates failure or saturation, not efficiency, in the presence of high output (symptoms). |
| **Regulatory effort** | Fasting insulin: the hormonal signal required to maintain glucose homeostasis. | Pain Compensatory System (PCS): the neurophysiological effort required to regulate pain. |
| **Functional outcome** | Fasting glucose: the systemic metabolic state being regulated. | Symptom burden: the clinical state of impairment (FIQR or reversed SF-36 PF). |
| **Formula structure** | $[\text{insulin} \times \text{glucose}] \div 22.5$ (normalised to a population constant). | $[\text{PCS} \times \text{symptoms}] \div \text{cohort mean}$ (normalised to a cohort average of 1.0); inputs are standardised scores and require a location shift that HOMA-IR does not. |
| **Biological impairment** | Chronic inflammation impairs insulin-receptor signalling (e.g., via IRS-1 phosphorylation). | Neuroinflammation impairs receptor sensitivity and inhibitory interneuron gain. |
| **State of resistance** | Metabolic resistance: the body is working harder (more insulin) to achieve less (higher glucose). | Cortical resistance: the brain is working harder (higher PCS) to achieve less (higher symptoms). |
| **Treatment goal** | Metabolic recalibration: lowering glucose while reducing insulin requirements. | Neurophysiological recalibration: lowering symptoms while reducing cortical effort (PCS). |

Note to Table 2. The CPRI addresses a paradox in chronic pain research: why the same neurophysiological signature can carry opposite clinical meaning in different patients. Just as an insulin concentration is healthy for one person and resistant for another depending on concurrent glucose, a high PCS signals cortical pain-compensatory resistance only when it fails to keep symptom burden low. Successful rehabilitation is defined by recalibration — restoration of a state in which the brain regulates pain efficiently with less effort.

We combined EEG theta asymmetry and intracortical inhibition (ICI) to derive a Pain Compensatory System (PCS) index and subsequently calculated CPRI as the mean-normalised product of PCS engagement and symptom burden. The resulting measure was designed to distinguish compensatory engagement from compensatory efficiency: a high CPRI represents substantial cortical compensatory engagement in the presence of persistent symptom burden and is therefore interpreted as a state of greater compensatory resistance or saturation, whereas a lower CPRI reflects a more favourable relationship between compensatory engagement and clinical pain control. Importantly, the index is not itself the mechanistic hypothesis; rather, it provides an operational measure with which to test that hypothesis. The central question is whether allostatic load modifies the clinical effectiveness of cortical compensation. We examined this model across two mechanistically distinct chronic pain cohorts — fibromyalgia, representing predominantly nociplastic pain, and knee osteoarthritis, representing predominantly structural/nociceptive pain— to determine whether the proposed relationship generalises across differing pain phenotypes and also whether rehabilitation alters that resistance state under high allostatic load.

## RESULTS

### Impaired Neurocompensatory Capacity Phenotype: Distribution and Validation

Participant characteristics by cohort and INCP status are summarised in Table 3. INCP groups did not differ in age or sex in FM; in OA, INCP+ participants were slightly younger (65.9 versus 72.4 years) with similar sex distribution and Kellgren–Lawrence grade, and, as expected by construction, INCP+ participants in both cohorts had higher BMI and higher depression, sleep and stress or anxiety scores. Allostatic load was operationalised as the Neurocompensatory Burden Index (NBI), a count of binary burden factors, and dichotomised as the Impaired Neurocompensatory Capacity Phenotype (INCP+ versus INCP−; Methods). In the fibromyalgia (FM) cohort, 56 of 116 participants (48.3%) were classified as INCP+ using a threshold of ≥3 of 5 NBI criteria. The NBI distribution was centred at scores of 2–3, with most participants (60.3%) exhibiting 2 or 3 burden factors, confirming meaningful heterogeneity without floor or ceiling effects. Only 4 participants (3.4%) had NBI = 0 and 2 (1.7%) met all five criteria. Sleep disturbance was the most prevalent component (87.9%), followed by perceived stress (54.3%), obesity (41.4%), depression (30.2%), and smoking (27.6%) (Figure 2). In the osteoarthritis (OA) cohort, using a 4-component NBI (body-mass index, depression, anxiety, sleep disturbance) with threshold ≥2, 35 of 65 neurophysiological subsample participants (53.8%) were classified as INCP+. Obesity (56.9%) and sleep disturbance (60.0%) were the dominant components in OA, contrasting with FM where sleep disturbance and perceived stress dominated, reflecting the mechanistically distinct allostatic load profiles of the two conditions (Figure 2).

**Figure 2.**
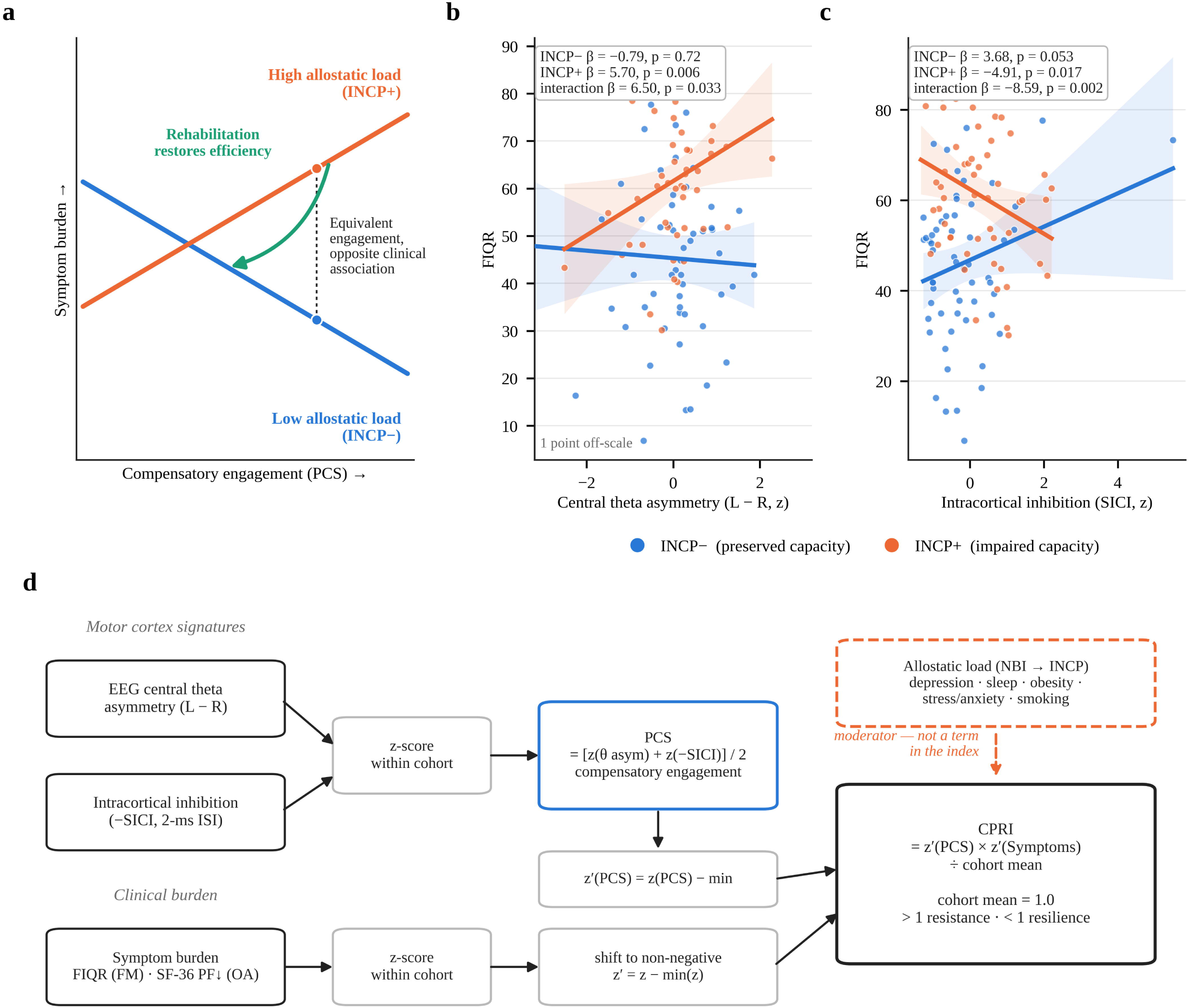
Distribution of neurocompensatory burden across cohorts. a) NBI-score distribution in fibromyalgia, with NBI ≥3 defining INCP+. b) Prevalence of individual burden components in fibromyalgia. c) NBI-score distribution in the osteoarthritis EEG–TMS subsample (n=65), with NBI ≥2 defining INCP+. d) Prevalence of individual burden components in osteoarthritis. Numbers above bars indicate participant counts. Blue denotes INCP− and orange denotes INCP+. NBI, Neurocompensatory Burden Index; INCP, Impaired Neurocompensatory Capacity Phenotype.

**Table 3.**
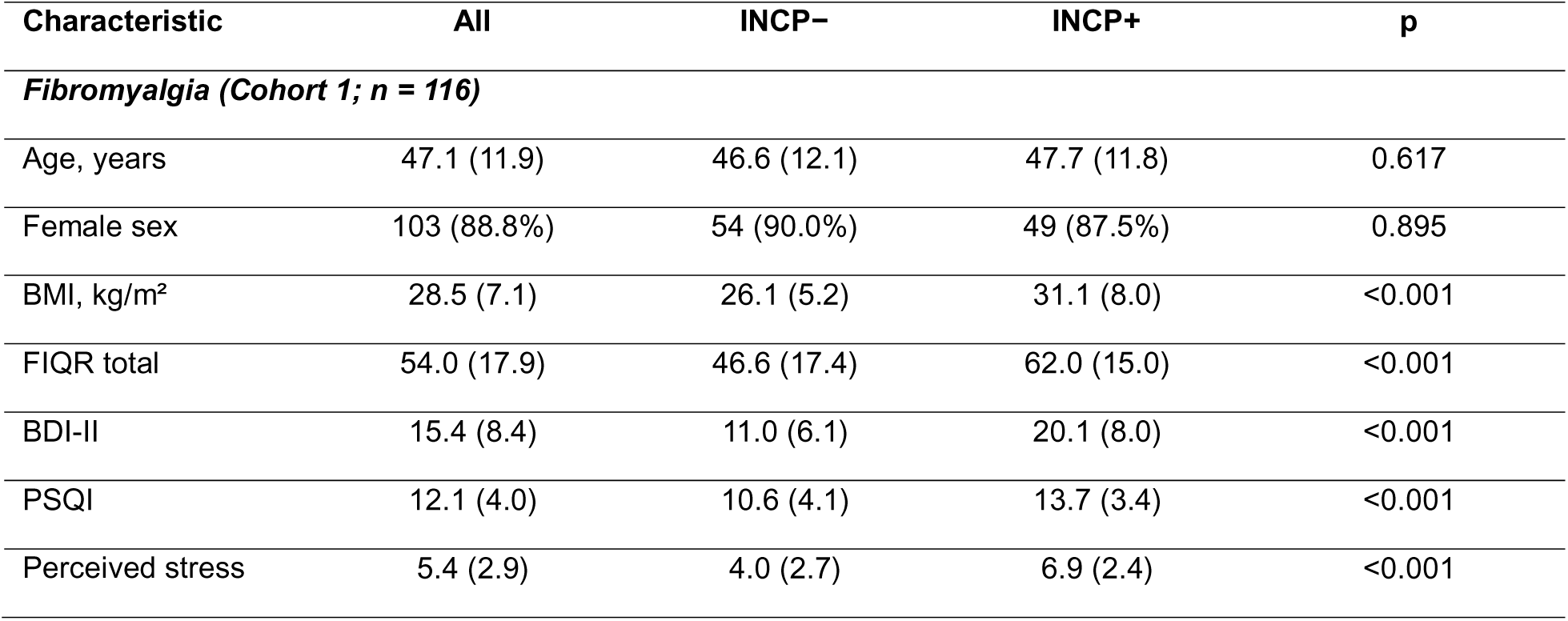
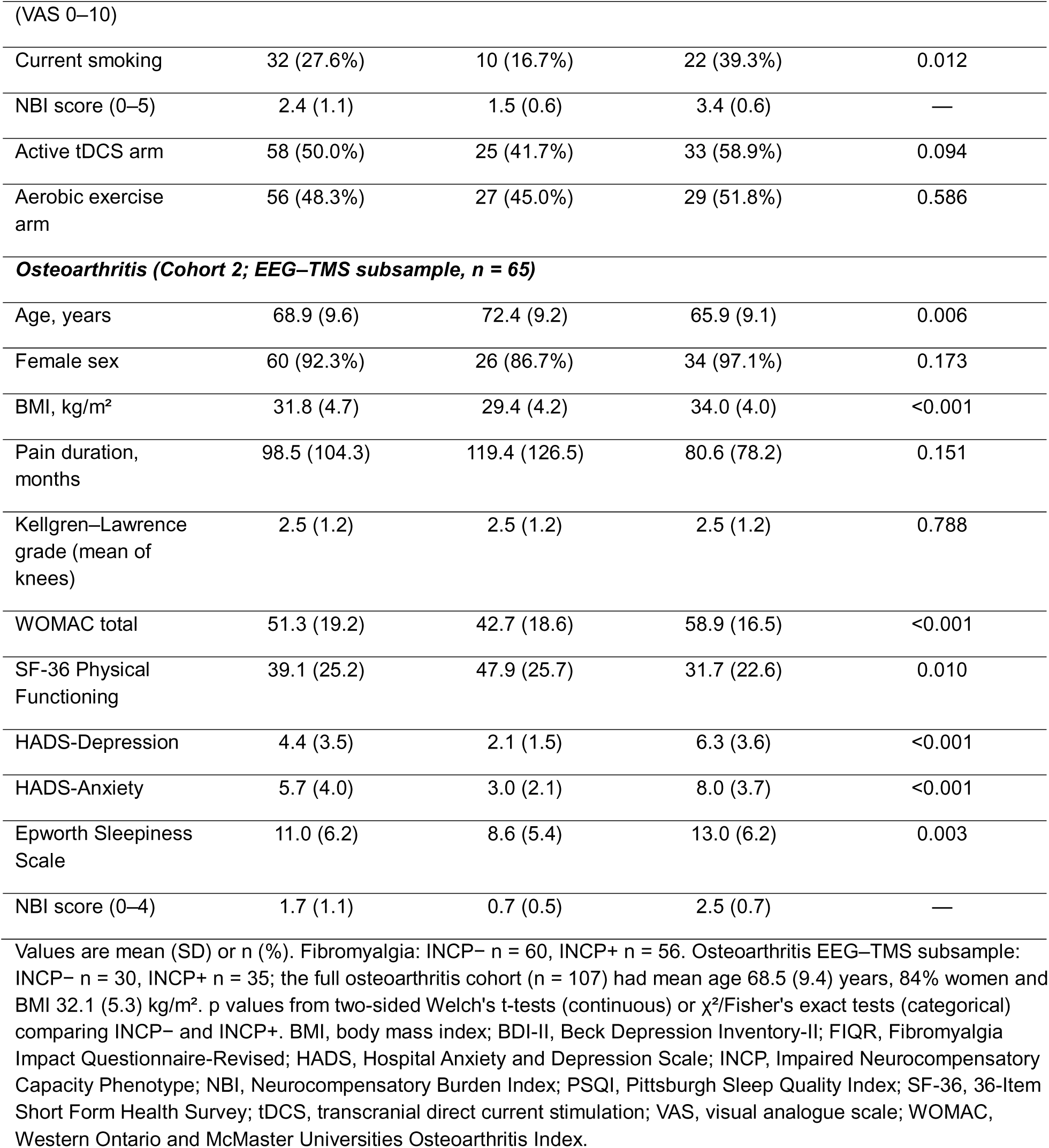
Participant characteristics by cohort and INCP status.

INCP status was strongly associated with symptom burden in both cohorts. In FM, INCP+ participants had substantially higher Fibromyalgia Impact Questionnaire–Revised (FIQR) scores than INCP− participants (M = 62.0 vs M = 46.6, d = 0.94; INCP main effect in all moderation models β = 17–28 points, p < 0.001). In OA, INCP+ participants showed significantly worse SF-36 Physical Functioning (SF-36 PF) than INCP− participants (M = 31.7 vs M = 47.9, p = 0.010, d = 0.64) and worse Western Ontario and McMaster Universities Osteoarthritis Index (WOMAC) total scores (M = 58.9 vs M = 42.7, p < 0.001), consistent across the full analytical sample (n = 107) and the neurophysiological subsample (n = 65). The pooled effect size for symptom burden by INCP status across both cohorts was d = 0.81 (95% CI [0.53, 1.08], p < 0.0001, I² = 0.0%), confirming that the INCP captures a robustly more impaired subgroup with complete consistency across mechanistically distinct chronic pain conditions.

### Neurophysiological Moderation by INCP

FM cohort — significant crossover interactions. Central theta asymmetry showed a significant crossover interaction with INCP status in predicting FIQR (β = 6.50, 95% CI [0.53, 12.46], p = 0.033, R² = 0.248). Among INCP− participants, theta asymmetry was not associated with FIQR (β = −0.79, p = 0.722); among INCP+ participants, greater left-lateralised theta activity was positively and significantly associated with worse symptoms (β = 5.70, 95% CI [1.70, 9.71], p = 0.006). Intracortical inhibition showed a parallel crossover interaction (β = −8.59, 95% CI [−14.08, −3.10], p = 0.002, R² = 0.245; Figure 1, panels b, c): among INCP− participants, higher inhibition trended toward lower FIQR (β = 3.68, p = 0.053); among INCP+ participants, higher inhibition was associated with higher FIQR (β = −4.91, p = 0.017) — the opposite pattern. The PCS Asymmetry–ICI composite showed the strongest interaction (β = 8.49, 95% CI [3.45, 13.52], p = 0.001, R² = 0.266), with higher PCS associated with lower symptoms in INCP− (β = −3.09, p = 0.096 trend) and higher symptoms in INCP+ (β = 5.40, p = 0.003). Specificity was confirmed: frontal alpha asymmetry showed no significant interactions, and the crossover pattern was selective to central theta, supporting the theoretical focus on M1 inhibitory dynamics.

OA cohort — directional replication. In the OA neurophysiological subsample (n = 65, INCP− = 30, INCP+ = 35), central theta asymmetry showed a directionally consistent crossover pattern against WOMAC Total: INCP− slope β = −0.16 (p = 0.970); INCP+ slope β = +2.10 (p = 0.442; WOMAC Pain INCP+ β = +1.13, p = 0.059 trend), a null association in INCP− and a positive but non-significant one in INCP+. The interaction did not reach significance (p = 0.659), given limited power with only 35 INCP+ participants. Bilateral short-interval intracortical inhibition (SICI) showed a significant protective association with WOMAC Total in INCP− participants (β = 7.29, SE = 3.31, p = 0.032) and WOMAC Pain (β = 1.34, SE = 0.51, p = 0.011), confirming that higher intracortical inhibitory tone associates with better symptom control in patients with preserved neurocompensatory reserve — replicating the FM directional finding. This protective association was systematically attenuated in INCP+ participants: the SICI slope for WOMAC Total in INCP+ was β = 3.15 (p = 0.272) — 43% of the INCP− magnitude — and for WOMAC Pain β = 0.70 (p = 0.229). The interaction terms did not reach significance (WOMAC Total: β = −4.14, p = 0.347; WOMAC Pain: β = −0.64, p = 0.404). Against SF-36 PF, the same attenuation pattern was observed for SICI — INCP− β = −9.67 (p = 0.001), INCP+ β = −3.88 (p = 0.312), retaining 40% of the INCP− magnitude— though interactions again did not reach significance given subsample power. This pattern — preserved ICI signal direction with attenuated magnitude in INCP+ — is distinct from the full crossover observed in FM and is consistent with a partial resistance state: in structurally-driven OA pain, the inhibitory system retains functional capacity and direction but its efficiency in translating inhibitory tone into symptom relief is degraded under high allostatic load.

### PCS Index Construction and Cross-Cohort Replication

The PCS Asymmetry–ICI index was constructed identically in both cohorts. Component intercorrelation was near-zero in both FM (r = 0.094, p = 0.346) and OA (r = 0.122, p = 0.334), confirming that theta asymmetry and intracortical inhibition capture independent, complementary neurophysiological dimensions — the defining property of a formative composite. Each component correlated with the composite at the value implied by their intercorrelation (FM: r = 0.732 and 0.747; OA: r = 0.749 for both), confirming correct index computations. The index exhibited a mean near zero and appropriate dispersion in both cohorts (FM: M = 0.00, SD = 0.74; OA: M = 0.00, SD = 0.75), consistent with expectations for an averaged z-score composite. Critically, PCS did not differ by INCP group at baseline in either cohort (FM: p = 0.271; OA: p = 0.248; Figure 3, panels c, d), consistent with allostatic load being associated with the efficiency of compensatory engagement rather than its magnitude. In FM, PCS was stable across the treatment period at the whole-group level (paired t, p = 0.616; no significant treatment-arm differences); as shown below, this group-level stability masked opposite changes in responders and non-responders.

**Figure 3.**
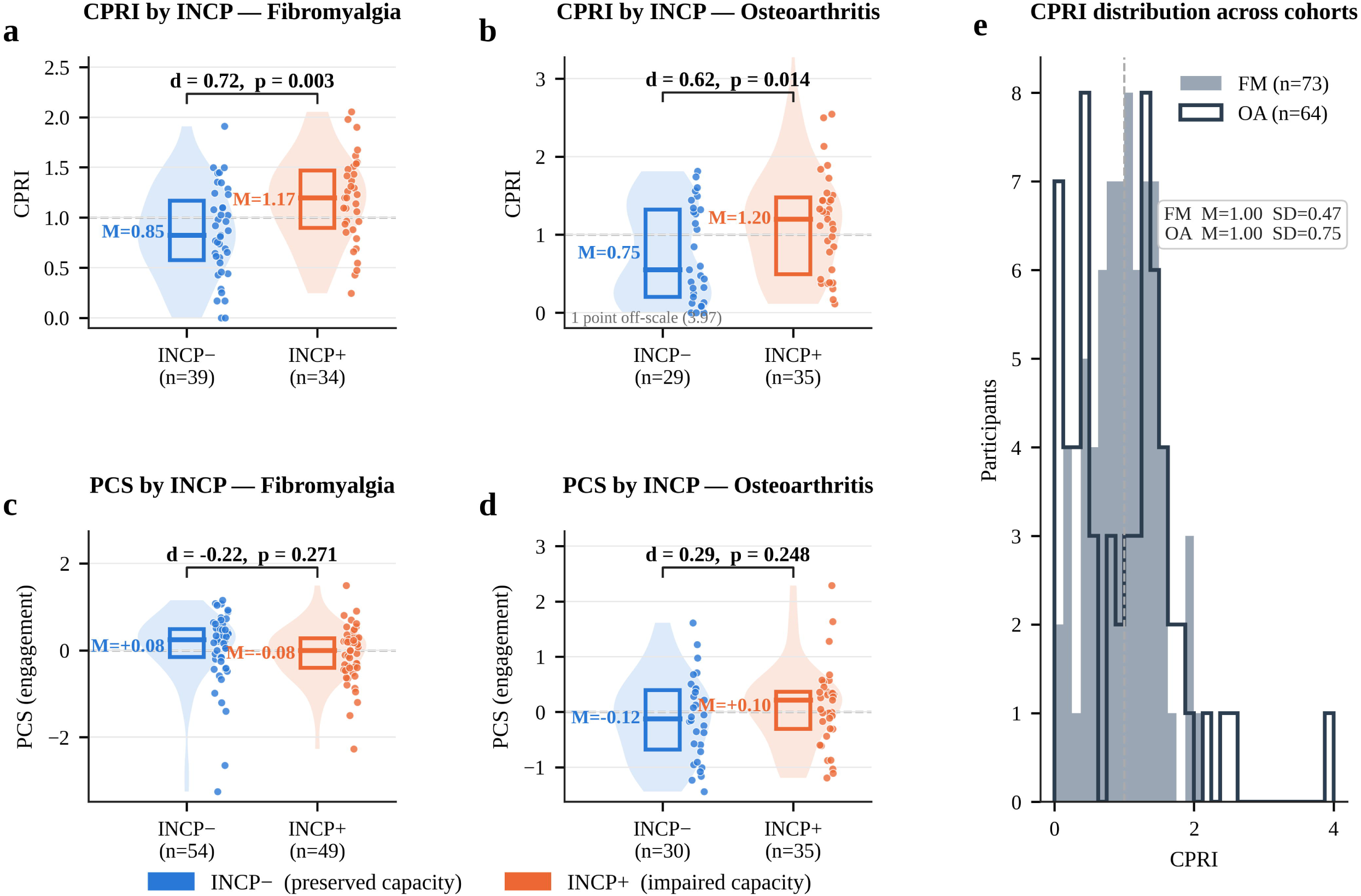
Cortical Pain-Compensatory Resistance Index and compensatory engagement according to allostatic load. CPRI distributions in INCP− and INCP+ participants with a) fibromyalgia and b) osteoarthritis; one osteoarthritis participant (CPRI 3.97) lies above the plotted range. c, d) The PCS index — compensatory engagement without the symptom term — in INCP− and INCP+ participants with c) fibromyalgia (n = 103) and d) osteoarthritis (n = 65): engagement does not differ by allostatic load in either cohort. e) CPRI distributions across the fibromyalgia and osteoarthritis cohorts; the dashed line indicates the normalised mean of 1.0. Group differences were assessed using two-sided Welch’s t-tests; brackets report Cohen’s d. Points represent participants, violins represent distributions, and boxes show medians and interquartile ranges. Blue denotes INCP− and orange denotes INCP+. CPRI, Cortical Pain-Compensatory Resistance Index; INCP, Impaired Neurocompensatory Capacity Phenotype; PCS, Pain Compensatory System.

### CPRI Distribution and Construct Validity

The CPRI was computed using complete pre- and post-treatment data, yielding analytical samples of 73 FM participants and 64 OA participants; of the 64 OA participants, 57 had post-treatment SF-36 PF (prognostic analyses) and 54 had both post-treatment neurophysiology and SF-36 PF (longitudinal analyses). In both cohorts the index was normalised to a population mean of 1.00 by construction; for the OA longitudinal analyses, standardisation and mean-normalisation were repeated within the 54-participant complete-data subsample. In FM, the CPRI exhibited a mean of 1.00 (SD = 0.47, range 0–2.05, skewness = −0.03). In OA (Figure 3, panel e), the CPRI — constructed using SF-36 PF as the symptom burden component (negated so that higher = greater functional impairment) — showed greater dispersion (M = 1.00, SD = 0.75) and mild positive skew (1.04), consistent with greater structural heterogeneity in OA where Kellgren–Lawrence grade introduces additional functional variance independent of neurophysiology (Figure 3, panels b, e). Construct validity was confirmed in both cohorts: CPRI correlated significantly with its PCS component (FM: r = 0.53, p < 0.001; OA: r = 0.78, p < 0.001) and with symptom burden (FM: r = 0.78, p < 0.001; OA: r = 0.62, p < 0.001), confirming that the index captures meaningful variance in both neurophysiological engagement and clinical burden without collapsing to either alone.

### CPRI is Elevated in Patients with High Allostatic Load

Consistent with the theoretical framework, CPRI was significantly elevated in INCP+ versus INCP− participants in both cohorts (Figure 3, panels a, b). In FM, INCP+ participants had mean CPRI of 1.17 (SD = 0.44) versus 0.85 (SD = 0.45) for INCP− (Welch t = 3.06, p = 0.003, d = 0.72). In OA, the same pattern was observed: INCP+ M = 1.20 (SD = 0.80) versus INCP− M = 0.75 (SD = 0.62; Welch t = 2.53, p = 0.014, d = 0.62). Fixed-effects meta-analysis yielded a pooled effect size of d = 0.67 (95% CI [0.33, 1.02], p = 0.00014), with directionally identical and numerically similar effects across cohorts (FM d = 0.72, OA d = 0.62; I² = 0.0%, noting that heterogeneity estimates are unreliable with k = 2 cohorts), indicating that CPRI elevation in individuals with high allostatic load is convergently observed across two conditions with fundamentally different pain mechanisms — consistent with cortical pain-compensatory resistance as a condition-agnostic property of the brain under allostatic load.

### CPRI Predicts Treatment Response and Tracks Rehabilitation Outcomes

Baseline CPRI predicted clinical improvement following rehabilitation in both cohorts. In FM, baseline CPRI correlated negatively with FIQR change (r = −0.47, p < 0.001, n = 73), indicating that greater CPRI at baseline was associated with greater symptom improvement. In OA, the association replicated in the expected direction (r = +0.50, p < 0.001, n = 57; positive sign reflects the SF-36 PF convention where higher change = improvement) and remained significant after adjustment for Kellgren–Lawrence grade and INCP status (β = 14.66, SE = 3.34, p < 0.001) (Figure 4). Notably, while baseline symptom severity showed the expected regression-to-the-mean association with improvement (FM: r = −0.27, p = 0.020; OA: r = −0.35, p = 0.007), the CPRI provided additional and stronger prognostic information: when baseline CPRI and baseline symptom severity were entered as competing predictors, CPRI remained associated with improvement in both cohorts (FM: β = −25.10, p = 0.0002; OA: p = 0.001), whereas baseline FIQR did not (p = 0.12). This reflects the distinct contribution of neurocompensatory state to treatment response — the index captures not how much pain the patient has, but how inefficiently the brain is regulating it.

**Figure 4.**
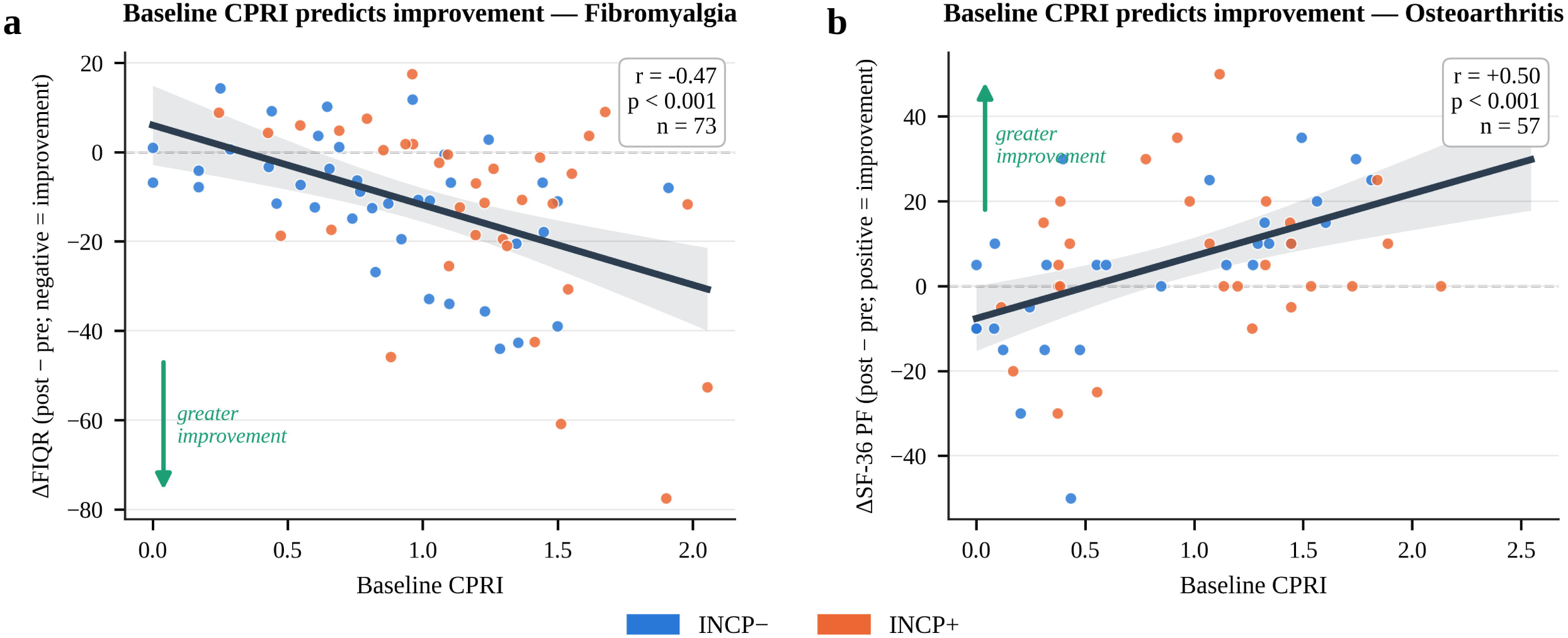
Baseline Cortical Pain-Compensatory Resistance Index and clinical improvement. Associations between baseline CPRI and clinical change in a) fibromyalgia and b) osteoarthritis (SF-36 Physical Functioning). More negative FIQR change and more positive SF-36 Physical Functioning change indicate greater improvement. Points represent participants, lines represent linear regression estimates, and shading indicates 95% confidence intervals. Pearson correlations and two-sided p values are displayed. Blue denotes INCP− and orange denotes INCP+. CPRI, Cortical Pain-Compensatory Resistance Index; FIQR, Fibromyalgia Impact Questionnaire-Revised; SF-36, 36-Item Short Form Health Survey.

The longitudinal CPRI trajectory by clinical response was consistent across cohorts. In FM, clinical responders (FIQR improvement ≥14 points; n = 23) showed a mean CPRI reduction of −0.85 units (SD = 0.43) following rehabilitation, while non-responders (n = 50) showed a mean change of +0.05 units (SD = 0.48; Welch t = 7.91, p < 0.001, d = −1.91) (Figure 5, panels a, b). This pattern replicated in OA: responders (SF-36 PF improvement ≥10 points; n = 25) showed CPRI reduction of −0.64 units (SD = 0.69), while non-responders (n = 29) showed CPRI increase of +0.17 units (SD = 0.70; Welch t = 4.30, p < 0.001, d = −1.17) (Figure 5, panels d, e). Critically, the neurophysiological PCS component — which contains no symptom term — moved with clinical response in both cohorts (Figure 5, panels c, f). In FM, ΔPCS was −0.35 (SD 0.87) in responders and +0.26 (SD 1.08) in non-responders (Welch t = −2.40, p = 0.020, d = −0.56); although PCS was stable at the whole-group level (paired t, p = 0.616), this reflected opposite changes in the two response groups rather than the absence of change. In OA, ΔPCS was −0.45 in responders and +0.10 in non-responders (p = 0.024). In both cohorts ΔPCS independently predicted ΔCPRI after controlling for symptom change (FM: β = 0.28, p < 0.001, controlling for ΔFIQR; OA: β = 0.665, p < 0.001, controlling for ΔSF-36 PF) — confirming that the CPRI trajectory reflects genuine neurophysiological recalibration, not merely symptom-driven regression to the mean. The pooled meta-analytic effect was d = −1.54 (95% CI [−1.95, −1.12], p < 0.0001, I² = 67.7%), with heterogeneity — not reliably estimable with two cohorts — reflecting the larger effect in FM (d = −1.91) than OA (d = −1.17), a gradient that would be consistent with the greater centrality of motor cortex inhibitory mechanisms in nociplastic pain, while the direction of CPRI reduction in responders versus increase in non-responders was identical across both conditions. In FM, these longitudinal associations were unchanged when the trial’s randomised factors were included as covariates (ΔCPRI by response β = −0.91 versus −0.90 unadjusted; ΔPCS by response β = −0.67, p = 0.012; baseline CPRI predicting ΔFIQR after adjustment for baseline FIQR and allocation β = −26.7, p < 0.001); neither tDCS nor exercise allocation was associated with any outcome (all p ≥ 0.09), and responders were distributed evenly across the four arms. These results support the proposition that successful rehabilitation restores neurocompensatory resilience — not merely suppresses symptoms: patients who improved achieved a recalibration of the motor cortex compensatory system requiring less cortical effort to achieve equivalent or better pain regulation.

**Figure 5.**
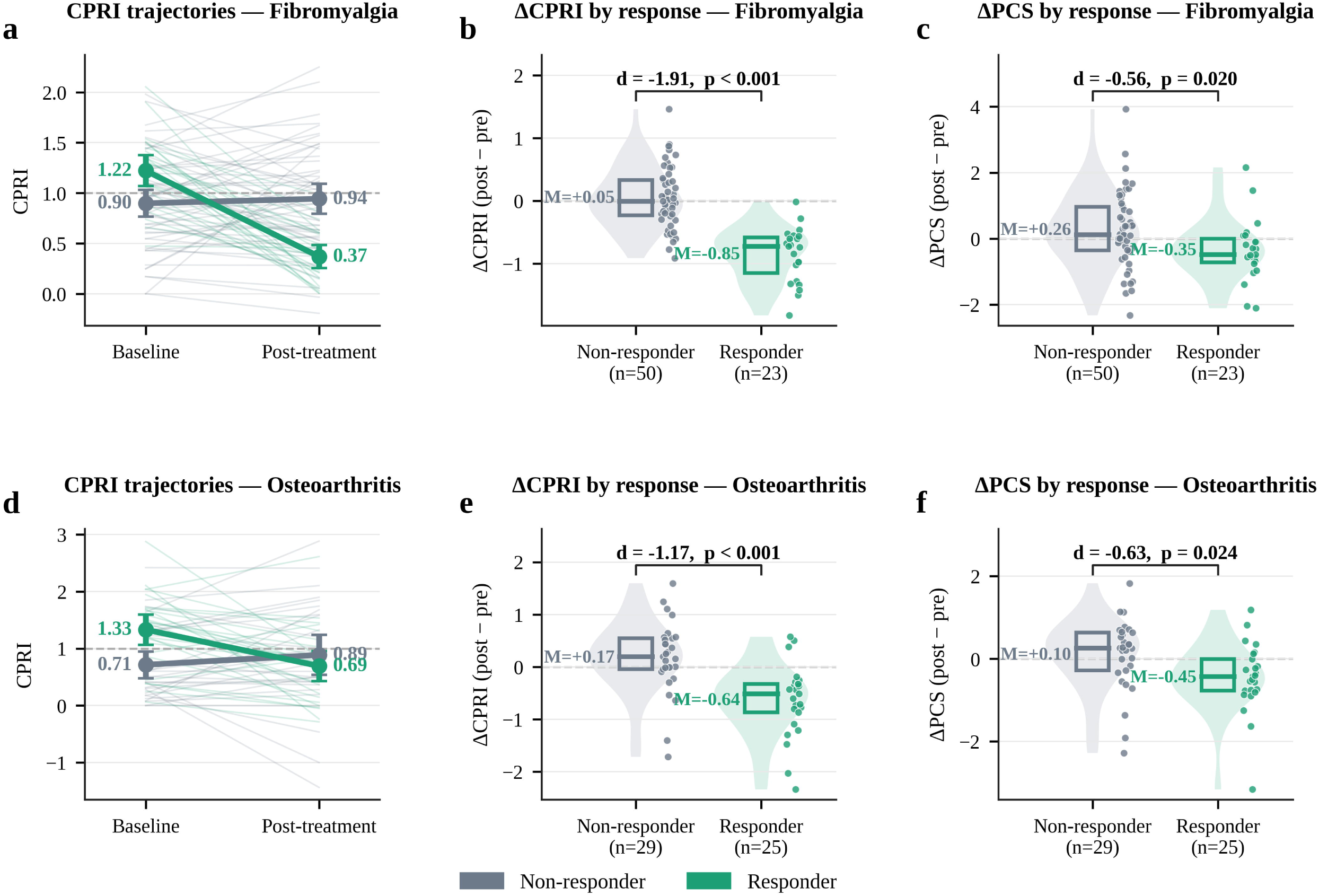
Cortical Pain-Compensatory Resistance Index and compensatory-engagement trajectories according to clinical response. a) Individual baseline-to-post-treatment CPRI trajectories in fibromyalgia responders and non-responders; thin lines represent individual participants and thick lines group means with 95% confidence intervals. b) Change in CPRI and c) change in the PCS index (post − pre) in fibromyalgia responders and non-responders. Response was defined as an FIQR improvement ≥ 14 points. d–f) The same for osteoarthritis, where response was defined as an SF-36 Physical Functioning improvement ≥ 10 points. Panels c and f show the neurophysiological component alone, which contains no symptom term: it fell in responders and rose in non-responders in both cohorts. Violins show distributions, boxes indicate medians and interquartile ranges, and points represent participants. Group differences were assessed using two-sided Welch’s t-tests; brackets report Cohen’s d. Grey denotes non-responders and green responders. CPRI, Cortical Pain-Compensatory Resistance Index; FIQR, Fibromyalgia Impact Questionnaire-Revised; PCS, Pain Compensatory System; SF-36, 36-Item Short Form Health Survey.

### Meta-analytic Synthesis

Fixed-effects meta-analysis pooled the four pre-specified estimates across cohorts (Figure 6). All four were directionally identical in both cohorts and all four reached p < 0.001; the baseline CPRI–improvement association pooled to d = −1.10 (95% CI −1.47 to −0.73). Heterogeneity was low except for the CPRI trajectory estimate (I² = 67.7%), where the larger FM effect would be consistent with the greater centrality of motor cortex mechanisms in nociplastic pain, although I² is not reliably estimable with two cohorts

## DISCUSSION

The principal finding of this study is that motor-cortical pain compensation appears to be a modifiable physiological state whose clinical effectiveness depends on cumulative allostatic load. The same neurophysiological features — central theta asymmetry and intracortical inhibition — had different associations with clinical status according to an individual’s psychological and metabolic load. In patients with relatively preserved compensatory reserve (INCP−), greater cortical inhibitory engagement was either unrelated or inversely related to symptom severity, consistent with effective homeostatic regulation. By contrast, under high allostatic load (INCP+), greater engagement was associated with greater symptom burden in fibromyalgia, and its protective association was attenuated in osteoarthritis, suggesting that increased cortical recruitment may coexist with diminished regulatory efficiency. Thus, the relevant distinction may not be whether the motor cortex engages compensatory mechanisms, but whether that engagement translates into effective pain control.

**Figure 6.**
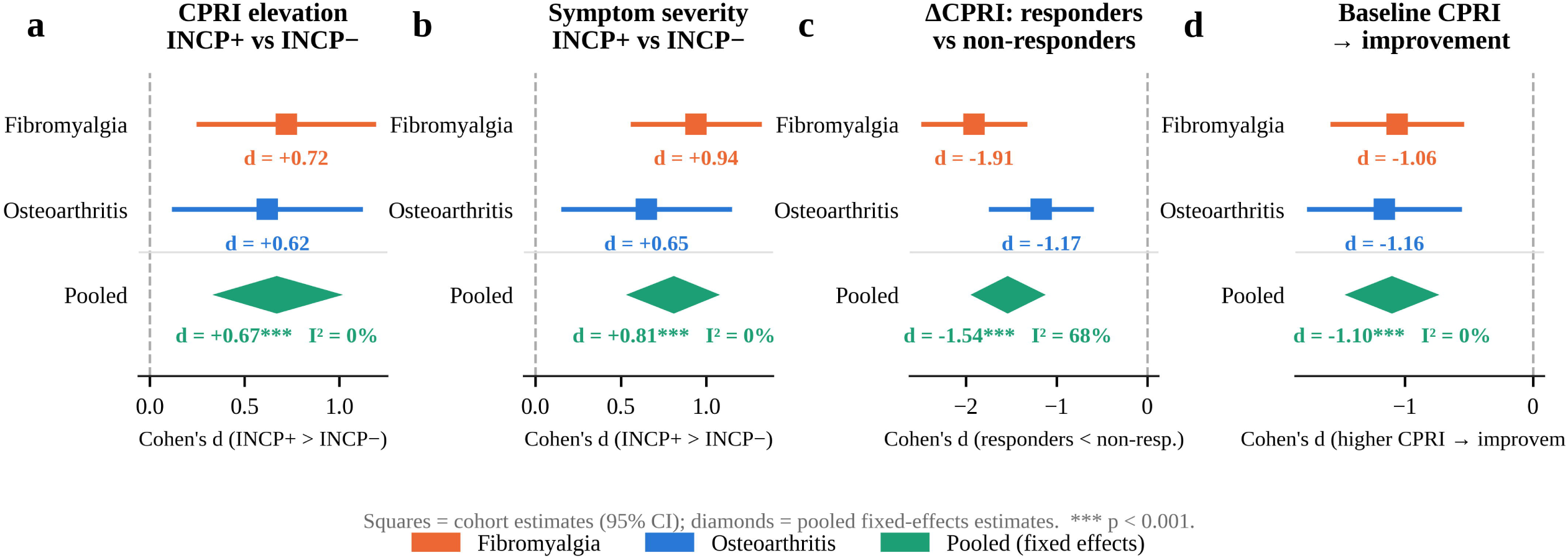
Cross-cohort synthesis of Cortical Pain-Compensatory Resistance Index findings. Cohort-specific and pooled effect sizes for a) CPRI by INCP status; b) symptom severity by INCP status; c) CPRI change in responders versus non-responders; and d) the association between baseline CPRI and improvement. Squares and horizontal lines represent cohort-specific Cohen’s d estimates and 95% confidence intervals. Diamonds represent inverse-variance-weighted fixed-effects estimates. Dashed lines indicate the null, and I² quantifies between-cohort heterogeneity. Orange denotes fibromyalgia, blue osteoarthritis, and green pooled estimates. CPRI, Cortical Pain-Compensatory Resistance Index; INCP, Impaired Neurocompensatory Capacity Phenotype.

To quantify this relationship, we developed the Cortical Pain-Compensatory Resistance Index (CPRI), a measure derived from motor-cortical inhibitory dynamics and conceptually modelled on the Homeostatic Model Assessment of Insulin Resistance (HOMA-IR; Table 2). CPRI was developed in fibromyalgia and subsequently evaluated in an independent osteoarthritis cohort with a mechanistically distinct pain phenotype. The conceptual parallel with insulin resistance is useful because it distinguishes compensatory engagement from compensatory efficiency: elevated insulin can accompany preserved glycaemic control when compensation remains effective, whereas the same response in the presence of hyperglycaemia indicates reduced regulatory efficiency. Similarly, a high CPRI represents substantial cortical compensatory engagement occurring in the presence of persistent symptom burden. Importantly, when baseline CPRI and baseline symptom severity were entered simultaneously as predictors, CPRI remained associated with subsequent improvement in both cohorts (FM, p = 0.0002; OA, p = 0.001), whereas baseline severity was not independently predictive in FM (p = 0.12). These findings suggest that compensatory state and clinical burden capture partially distinct dimensions of the pain-regulatory system.

A reasonable objection is that neither central theta asymmetry nor intracortical inhibition has been established as a compensatory signal, and we do not claim to have proved that here. We instead propose three criteria for judging a putative compensatory signature and note how the present data bear on each. First, a compensatory signal should be engaged across the spectrum of challenge rather than exhausted in the most affected patients: in both cohorts, PCS did not differ by allostatic load. Second, its engagement should accompany better clinical status when regulatory reserve is preserved: in INCP− participants, stronger inhibition and greater left-lateralised theta were unrelated or inversely related to symptoms, and in osteoarthritis the inhibitory association was protective. Third, it should recalibrate with recovery rather than remain fixed: the neurophysiological component moved with clinical response in both cohorts, independently of symptom change. These criteria mirror those used in the cognitive-reserve literature, where additional neural activity is judged compensatory by its relationship to outcome under differing levels of pathology rather than by its magnitude alone.(8, 9) The theta component remains the less established of the two: its lateralisation was specified a priori, and its crossover was clear in fibromyalgia but only directionally consistent in osteoarthritis. Whether central theta reflects thalamo-cortical dysrhythmia, a sensorimotor idling rhythm or compensatory recruitment of M1 cannot be resolved by this design; what the data show is that its clinical meaning depends on load, as a compensatory mechanism would predict. The contribution of the present work is therefore less the specific signals than the framework: motor cortex pain modulation characterised in terms of reserve and resistance, in which the same signal can indicate effective compensation or compensatory saturation depending on the load against which it operates.

This framework has parallels with other biological systems in which the magnitude of a compensatory response cannot be interpreted independently of the load against which it operates. In neurodegenerative disease, reserve-related neural activity modifies the relationship between pathological burden and cognitive performance: individuals with greater reserve can maintain function despite comparable pathology,(8) while compensatory processes may intensify with accumulating pathology before declining as reserve becomes exhausted.(9) A similar principle may apply here. PCS engagement did not differ according to INCP status in either cohort, suggesting that allostatic load primarily modified the effectiveness of cortical compensation rather than its overall magnitude. In metabolic physiology, autonomic state similarly influences the efficiency of pancreatic compensation: lower heart-rate variability is associated with a reduced composite insulin response to glucose challenge across the spectrum of glucose tolerance.(24) Together, these observations support a broader regulatory principle in which the physiological significance of a compensatory signal depends jointly on the magnitude of the response and the systemic environment in which it occurs.

The biological pathways through which cumulative allostatic load could alter cortical compensatory efficiency remain to be established. One plausible mechanism is microglia-mediated neuroinflammation. Components of the Neurocompensatory Burden Index — including depression, sleep disturbance, obesity and anxiety — have been linked to inflammatory signalling through partially overlapping mechanisms — HPA-axis dysregulation, microglial priming, adipokine signalling and altered glucocorticoid-receptor function.(13, 17, 25–28) Microglia-derived TNF-α can reduce GABA_A-receptor surface expression and impair inhibitory neurotransmission,(29) and TSPO PET studies have demonstrated evidence of glial activation in fibromyalgia.(30) These observations provide a mechanistic basis for the hypothesis that systemic allostatic load could impair the efficiency of GABAergic inhibitory processes indexed by SICI. This pathway remains speculative, however: to our knowledge, cortical glial activation has not been directly linked to intracortical inhibition in humans, although intracortical inhibition itself is consistently associated with symptom burden in these populations, with the direction of the association varying by condition and compensatory state.(31, 32) Testing whether inflammatory measures such as CRP, IL-6 or PET-derived indices of glial activation mediate the relationship between allostatic load and CPRI would provide a direct test of this mechanism.

An important feature of the present findings is their replication across two pain conditions with different predominant mechanisms. CPRI showed directionally consistent associations in fibromyalgia and osteoarthritis despite differences in clinical phenotype, assessment instruments and geographical setting. This convergence suggests that cortical pain-compensatory resistance may reflect a regulatory property that extends beyond a single diagnostic category, consistent with the position of M1 as a hub for rehabilitation-induced analgesia,(1) although inference regarding between-condition generalisability remains limited by the inclusion of only two cohorts. The moderation patterns were also not identical. Fibromyalgia showed a crossover pattern in which the relationship between inhibitory engagement and clinical status changed direction, whereas osteoarthritis showed predominantly attenuation rather than reversal. One possible explanation is that structural nociceptive input contributes independent variance to symptoms in osteoarthritis, thereby constraining the extent to which systemic load can determine the relationship between cortical regulation and clinical expression. This interpretation should be tested directly rather than inferred from the present data.

The longitudinal findings further suggest that compensatory resistance is dynamic rather than trait-like. CPRI decreased among treatment responders in both cohorts and did not decrease among non-responders (unchanged in fibromyalgia, increased in osteoarthritis). Importantly, this pattern was not explained solely by the symptom component incorporated into the index. In fibromyalgia, baseline CPRI remained associated with improvement after adjustment for baseline FIQR (β = −25.10, p = 0.0002), whereas baseline FIQR itself was no longer independently associated with outcome (p = 0.12). In osteoarthritis, CPRI similarly remained associated with outcome after adjustment for baseline SF-36 physical functioning (p = 0.001). Moreover, the neurophysiological PCS component — which contains no symptom term — changed more among responders than non-responders in both cohorts (fibromyalgia ΔPCS −0.35 versus +0.26, p = 0.020; osteoarthritis −0.45 versus +0.10, p = 0.024), and change in PCS remained associated with change in CPRI after adjustment for symptom change in each cohort. In fibromyalgia, PCS was stable at the whole-group level (p = 0.616), but this reflected opposite movements in responders and non-responders rather than the absence of neurophysiological change. These analyses argue against a purely arithmetic explanation in which the prognostic behaviour of CPRI arises from its inclusion of baseline symptom severity, and instead support a contribution from longitudinal change in the underlying neurophysiological component.

Several steps are required before CPRI can be considered for clinical application. Because the index is normalised to the sample mean, its absolute magnitude is inherently reference-dependent. Application across centres will therefore require either externally derived reference distributions or standardised acquisition and normalisation procedures, analogous to the calibration requirements of metabolic resistance indices. Preliminary response thresholds, distributional anchors and estimates of minimum important change are provided in Supplementary Note 1, but require prospective validation. Practical implementation also remains challenging because the present formulation requires multichannel EEG and paired-pulse TMS. Whether comparable information can be retained using reduced EEG montages or simplified neurophysiological protocols will be important for determining its eventual scalability.

The findings may also have implications for the interpretation of comorbidity in chronic pain. Depression, sleep disturbance, obesity and anxiety are conventionally treated as covariates or competing explanations for variation in pain outcomes. Our results instead raise the possibility that their cumulative load contributes to the physiological context in which cortical pain-regulatory mechanisms operate. Under this model, these factors need not simply confound associations between cortical physiology and pain; they may modify the efficiency with which cortical compensatory processes influence clinical outcomes. This distinction has therapeutic implications because interventions directed at systemic load could, in principle, alter the effectiveness of cortical regulation even without directly targeting M1. Prospective interventional studies will be required to establish whether modifying allostatic load produces corresponding changes in cortical compensatory efficiency.

This study has some limitations. Both cohorts were analysed retrospectively as secondary mechanistic analyses of existing datasets, and the smaller osteoarthritis neurophysiological sample (n = 65) limited power for moderation analyses. The operational definition of INCP also differed modestly between cohorts: smoking and perceived stress in fibromyalgia were replaced by anxiety in osteoarthritis because of differences in available measures. Although this heterogeneity provides some indication that the findings are not dependent on an identical set of variables, it also limits direct comparison and emphasises the need for a prospectively specified allostatic-load construct. Complete-case analyses were used throughout and may have reduced generalisability. Conversely, imputation of complex EEG and paired-pulse TMS measures in datasets of this size would require assumptions that are difficult to justify. Finally, cohorts combining longitudinal clinical outcomes with both multichannel EEG and paired-pulse TMS remain uncommon. Larger, prospectively designed studies incorporating standardised measures of cortical physiology, systemic load and inflammatory biology will therefore be required to validate the proposed model and establish its generalisability across chronic pain conditions.

Collectively, these findings support a model in which chronic pain is not necessarily characterised by failure to engage inhibitory compensatory mechanisms. Rather, cumulative allostatic load may alter the efficiency with which such engagement is translated into pain control. Under relatively preserved reserve, cortical inhibitory activity may accompany successful regulation; under greater systemic load, similar or greater engagement may coexist with persistent symptoms, consistent with a state of compensatory resistance. The analogy to insulin resistance is therefore mechanistic rather than merely descriptive: in both settings, the compensatory process remains engaged, but its capacity to restore homeostasis is reduced. The replication of this pattern across fibromyalgia and osteoarthritis, together with longitudinal changes in the neurophysiological component during rehabilitation, identifies compensatory efficiency — rather than compensatory engagement alone — as a potentially important dimension of chronic pain biology and a candidate target for patient stratification and treatment monitoring.

## METHODS

### Study design and parent cohorts

This study reports a mechanistic secondary analysis of two studies: a fibromyalgia (FM) randomised clinical trial (Cohort 1) and a knee osteoarthritis (OA) rehabilitation cohort (Cohort 2). Both cohorts are described in full elsewhere.(33, 34) All participants in both cohorts provided written informed consent, and both studies were conducted in accordance with the Declaration of Helsinki.

Study 1 — Fibromyalgia. Adults with FM were recruited through outpatient pain and rehabilitation clinics. Inclusion required confirmed FM, FIQR ≥ 40, and ability to undergo neurophysiological assessments. Exclusion criteria included unstable medical or psychiatric conditions and contraindications to TMS or EEG.(33, 35) Participants were randomly assigned to active or sham tDCS with aerobic or non-aerobic exercise. Assessments were conducted at baseline and post-treatment by blinded evaluators. The trial was registered (NCT03371225) and approved by the Mass General Brigham IRB (protocol 2017P002524).

Study 2 — Osteoarthritis. Adults with confirmed knee OA were recruited through outpatient rehabilitation clinics at the Institute of Physical Medicine and Rehabilitation, Hospital das Clínicas, University of São Paulo (IMREA/DEFINE cohort).(34, 36) Inclusion required clinical and radiographic confirmation of knee OA, age ≥ 50 years, and clinical stability. The individualised multimodal rehabilitation programme comprised neuromuscular electrical stimulation, focal extracorporeal shockwave therapy, radial pressure waves, and paraspinous lidocaine blocks, administered according to individual clinical findings over a median of 5.9 weeks. Clinical, functional, and neurophysiological assessments were conducted at baseline and post-rehabilitation. The study was approved by the Hospital das Clínicas Ethics Committee (CAAE: 86832518.7.0000.0068).

### Neurophysiological and clinical assessments

Both cohorts underwent EEG and TMS assessments by the same collaborative team following closely parallel acquisition protocols targeting the primary motor cortex (M1), enabling direct comparison of derived neurophysiological indices across conditions.

Clinical outcomes. Clinical outcomes were assessed using the Fibromyalgia Impact Questionnaire–Revised (FIQR) in FM and the SF-36 Physical Functioning subscale (SF-36 PF; range 0–100, higher = better functional capacity) in OA. SF-36 PF was selected as the primary OA outcome rather than a joint-specific pain index because it captures multidimensional functional disability across domains including walking, stair climbing, and activities of daily living, providing a measure conceptually parallel to the FIQR.(37) The WOMAC total score (72-hour recall) was retained as a secondary cross-sectional outcome for phenotype validation, where its shorter recall window and joint-specific nociceptive focus suit baseline characterisation of the allostatic load phenotype but are less suited to the broader functional burden that motivates the prognostic analyses. Clinical response was defined as ≥ 14-point FIQR improvement in FM and ≥ 10-point SF-36 PF improvement in OA, thresholds specified a priori as conservative, distribution-based response criteria (Supplementary Note 1).

Allostatic load component instruments. Depressive symptoms were assessed with the Beck Depression Inventory-II (BDI-II; ≥ 20 = moderate-to-severe depression per the standard severity classification)(38) in FM and the Hospital Anxiety and Depression Scale depression subscale (HADS-D; ≥ 8 = caseness)(39, 40) in OA. Sleep disturbance was assessed with the Pittsburgh Sleep Quality Index (PSQI; ≥ 8 = clinically significant sleep disturbance)(41) in FM and the Epworth Sleepiness Scale (ESS; ≥ 10 = excessive daytime sleepiness)(42) in OA. Perceived stress was assessed with a Visual Analogue Scale (VAS stress; 0–10; ≥ 6 = clinically meaningful burden) in FM. Anxiety was assessed with the HADS anxiety subscale (HADS-A; ≥ 8 = caseness)(39, 40) in OA. Obesity was defined as BMI ≥ 30 kg/m² in both cohorts per WHO criteria.(43) Smoking status was recorded as a binary variable (current smoker yes/no) in FM only, included as a burden indicator given its association with worse pain, sleep and mood outcomes in chronic pain.(44) Thresholds correspond to published cutoffs for each instrument; the PSQI threshold of ≥ 8 follows the higher cutoff recommended for clinical populations rather than the original > 5.(41, 45)

Transcranial magnetic stimulation (TMS). Cortical excitability and inhibition were assessed with single- and paired-pulse TMS over bilateral M1 (Magstim Rapid² system, figure-of-eight coil). Resting motor threshold (rMT) was the minimum intensity eliciting MEPs ≥ 100 µV in ≥ 3 of 5 trials. Short-interval intracortical inhibition (SICI; ICI analogue) was assessed at 2-ms interstimulus interval (conditioning stimulus 80% rMT; test stimulus 120% rMT; 10 trials/condition). SICI was the conditioned-to-unconditioned MEP amplitude ratio; lower values indicate stronger inhibition. SICI was assessed over both hemispheres as in the parent studies;(2, 33) bilateral SICI was computed for the present analyses as the mean of the two.

Electroencephalography (EEG). Resting-state EEG was recorded with 64-channel systems positioned per the international 10–20 system. Eyes-closed recordings (5 minutes; 4 analysed) were preprocessed in both cohorts with a standardised, previously validated MATLAB pipeline (filtering 1–90 Hz, 60-Hz notch, downsampling to 250 Hz, average re-reference, ICA artefact removal), ensuring identical preprocessing across cohorts.(46) Spectral power was computed using fast Fourier transformation. Relative power (%) was band power divided by total broadband power. Central electrodes (C3, C4, Cz) indexed M1-proximal oscillatory dynamics. EEG theta asymmetry was Left − Right central relative theta power (4–7.9 Hz); positive values reflect greater left-hemispheric theta engagement.

### Construction of the Impaired Neurocompensatory Capacity phenotype (INCP)

The INCP (Table 1) operationalises cumulative allostatic load — the physiological, metabolic, and psychological load that degrades neurocompensatory reserve. This framework is grounded in allostatic load theory, describing cumulative “wear and tear” when sustained stressors exceed adaptive capacity.(10, 47, 48) The Neurocompensatory Burden Index (NBI) domains are active stressors that converge on pain-modulatory circuits. Depression, anxiety, and sustained perceived stress dysregulate the hypothalamic–pituitary–adrenal (HPA) axis and activate pro-inflammatory cytokine cascades, impairing descending inhibitory control and cortical GABAergic tone.(16, 49) Sleep disturbance disrupts the synaptic homeostasis that sleep normally restores,(50) and, together with severe stress, acts as a glial activator that increases central excitability and degrades the balance between excitation and inhibition in chronic pain.(15, 51) Obesity promotes neuroinflammation through adipokine and cytokine signalling, blunting pain modulation and descending inhibition.(14, 17) Smoking, captured in the FM cohort only, is associated with worse pain, function, sleep and mood in large chronic pain registries.(44) Collectively, convergence on neuroendocrine, immune, and autonomic pathways impairs compensatory reserve, explaining why cumulative load determines the tipping point into cortical pain-compensatory resistance. Because depression, anxiety, and sleep disturbance constitute three of the five FM components and three of the four OA components, the INCP is as much a mental health construct as a pain construct: it captures the interface between psychological load and cortical neurophysiology that is central to the present framework.

FM cohort NBI — 5 components, threshold ≥ 3. Smoking (current, self-report); obesity (BMI ≥ 30 kg/m²); sleep disturbance (PSQI ≥ 8); perceived stress (VAS ≥ 6/10); and depressive symptoms (BDI ≥ 20). Each criterion was binary; NBI = sum of present criteria (range 0–5). INCP was assigned for NBI ≥ 3, yielding phenotype prevalence of 48.3%.

OA cohort NBI — 4 components, threshold ≥ 2. Obesity (BMI ≥ 30 kg/m²); depression (HADS-D ≥ 8); anxiety (HADS-A ≥ 8); and sleep disturbance (ESS ≥ 10). NBI = sum of present criteria (range 0–4). INCP was assigned for NBI ≥ 2. Smoking and perceived stress were unavailable in the OA cohort; anxiety replaced them based on its HPA-axis role and its association with altered intracortical inhibition in chronic pain populations.(31, 52) The classification thresholds (≥ 3 of 5 in FM; ≥ 2 of 4 in OA) were selected to identify participants with convergent multi-domain load without requiring exhaustive comorbidity, consistent with count-based operationalisations of allostatic load,(47) and reflected data availability while preserving the core construct logic.

### Construction of the PCS index

Rationale for M1 as the primary network node. The primary motor cortex was selected as the target node because it is a critical hub in the pain connectome. Evidence shows that M1 mediates analgesia through descending inhibitory pathways, thalamo-cortical modulation, cortico-limbic pain signature normalisation, and the endogenous opioid system.(1) M1 is embedded in cortico-limbic circuitry — anterior cingulate cortex, insula, amygdala and periaqueductal grey — through which affective states including depression, anxiety and stress shape nociceptive processing,(20, 53) and integrates these cognitive, affective and sensory signals with its descending analgesic output.(1)

Rationale for component selection. Two M1 signatures were selected for the PCS index, each hypothesised to index compensatory engagement rather than assumed to. First, intracortical inhibition (ICI), measured via SICI at 2-ms interstimulus interval, indexes GABAergic interneuron function within M1. A meta-analysis of TMS studies in fibromyalgia shows that reduced intracortical inhibition is a consistent neurophysiological signature of the condition, and that treatment-related increases in inhibition track pain reduction.(32) In patients with preserved neurocompensatory reserve, high SICI is proposed to reflect efficient M1 inhibitory engagement; in those with high allostatic load, the same SICI level is proposed to signal compensatory saturation.(2, 49, 54) Second, EEG theta asymmetry (Left − Right) at central electrodes overlying M1 was selected because theta oscillations in this region have been characterised by our group across fibromyalgia, osteoarthritis, and neuropathic pain cohorts as a context-dependent marker of adaptive neural engagement under chronic pain load.(4, 21, 22, 36) Left-hemisphere lateralisation was specified a priori rather than derived from these data, on two grounds: our prior characterisation of central theta activity as a compensatory signature,(4, 21, 22) and the prevailing convention of targeting the left hemisphere in prefrontal stimulation studies of pain. Independent support comes from a report of increased left centro-parietal theta coherence in fibromyalgia relative to pain-free controls.(55) Hemispheric lateralisation in pain processing is, however, not resolved: left dominance has been reported for the sensory-discriminative dimension of nociception,(23) whereas the amygdala literature describes right-lateralised pain-related plasticity,(56) and the left-target convention in prefrontal stimulation derives in part from depression protocols rather than from pain physiology. The direction of the asymmetry term was therefore fixed before analysis rather than selected on the basis of model fit.

Index construction (Figure 1). EEG theta asymmetry (Left − Right) and reverse-coded SICI (−SICI; higher = stronger inhibition) were each z-scored within their respective cohorts and averaged:

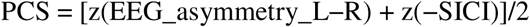

Higher PCS values indicate greater left-lateralised theta activity combined with stronger intracortical inhibition — the coordinated M1 inhibitory signature. The index was computed only for participants with non-missing data on both components (FM: n = 103; OA EEG+TMS subsample: n = 65). The PCS is a formative composite: unlike reflective indices, whose components are expected to intercorrelate and whose validation target is internal consistency,(57) theta asymmetry and intracortical inhibition index complementary neurophysiological mechanisms rather than a single underlying latent trait, so the near-zero intercorrelation observed here is the expected property rather than a psychometric failure (Figure 1). The index was therefore validated against external criteria rather than by internal consistency.

### Construction of the Cortical Pain-Compensatory Resistance Index (CPRI)

Following the form of HOMA-IR,(18, 19) the CPRI (Figure 1 and Table 2) quantifies the mismatch between cortical compensatory effort (PCS) and symptom burden — operationalised as FIQR in FM and SF-36 PF (reversed so that higher = greater functional impairment) in OA. Both components were z-scored and shifted to non-negativity by subtracting the minimum value in the respective cohort — a step HOMA-IR does not require, because its inputs are ratio-scaled assay values rather than standardised scores (Supplementary Note 1) — and their product normalised by the sample mean:

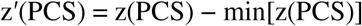

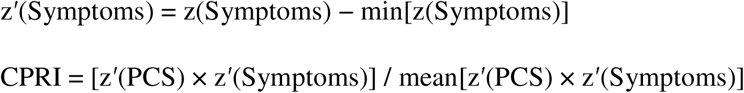

CPRI equal to 1.0 represents population-average compensatory–symptom mismatch. Post-treatment CPRI was computed from post-treatment PCS and symptom scores using the standardisation, shift and normalisation constants estimated at baseline, applied unchanged, so that post-treatment values are on the baseline scale.

### Statistical analysis

All analyses used a complete-case approach. Primary outcomes were FIQR total score in FM and SF-36 PF in OA (reversed for CPRI construction so that higher = greater impairment). In OA, WOMAC (72-hour recall) was used for cross-sectional moderation and SF-36 PF (4-week recall) for CPRI construction and prognostic analyses.

Moderation analyses. To test whether INCP status moderated the associations between neurophysiological measures and symptom severity, we fitted ordinary least squares (OLS) regression models with interaction terms, each of the general form:

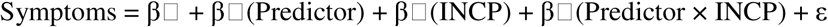

where the predictor was central theta asymmetry, the PCS composite, or bilateral SICI. All predictors were z-scored within cohort, so coefficients are per standard deviation of the predictor. In moderation models SICI entered as the standardised conditioned-to-unconditioned ratio (lower = stronger inhibition), so a positive coefficient denotes weaker inhibition associated with higher symptoms; the PCS uses reverse-coded SICI. The interaction coefficient β₃ was the primary parameter of interest, testing whether the slope relating the predictor to symptom severity differed between INCP groups; a significant interaction indicates that the strength or direction of the predictor–symptom relationship varies with neurocompensatory capacity, the core hypothesis of this study. In FM, models tested central theta asymmetry, the PCS composite, and SICI against FIQR; following a significant interaction, simple slopes were computed separately for each INCP group. As a specificity control, an identically specified model was fitted with frontal alpha asymmetry as the predictor, to confirm that the crossover pattern was selective to central theta rather than reflecting a global oscillatory effect. In OA, models tested central theta asymmetry and bilateral SICI against WOMAC total (72-hour recall) and SF-36 PF (4-week recall); prognostic analyses additionally adjusted for Kellgren–Lawrence grade and INCP status.

CPRI properties and meta-analysis. CPRI properties were evaluated using Pearson correlations, Welch t-tests, and Cohen’s d. Incremental validity was tested by entering CPRI alongside baseline symptom severity as competing predictors of outcome change. Anti-circularity was tested by examining whether the PCS neurophysiological component independently predicted ΔCPRI beyond symptom change alone, and whether ΔPCS itself differed by clinical response. A pre-specified fixed-effects meta-analysis with inverse-variance weighting pooled four estimates across cohorts: (i) CPRI elevation by INCP status; (ii) ΔCPRI by clinical response; (iii) symptom severity by INCP status; and (iv) baseline CPRI–improvement association. Effect sizes were Cohen’s d; for correlations, d = 2r/√(1−r²). Heterogeneity was assessed using I², noting that heterogeneity estimates are unreliable with k = 2 cohorts.

Statistical inference. Because neurophysiological indices and questionnaire-based outcomes can exhibit heteroskedasticity and sensitivity to influential observations, all inference used heteroskedasticity-consistent robust standard errors with the HC3 estimator, which outperforms HC0 and HC1 in finite samples with high-leverage observations.(58) Model results are reported as unstandardised regression coefficients (β) with HC3 robust standard errors, t statistics, two-sided p values, and 95% confidence intervals; model fit is summarised with R² and adjusted R², and overall model significance with the F statistic. Statistical significance was set at α = 0.05 two-sided; FDR q < 0.05 for families of EEG interaction tests. As a sensitivity analysis, the FM longitudinal models (ΔCPRI and ΔPCS by response; baseline CPRI predicting ΔFIQR) were refitted with the trial’s randomised factors (active versus sham tDCS; aerobic versus non-aerobic exercise) as covariates.

Software. Analyses were implemented in Python 3 (scipy, statsmodels), and cross-validated and checked in Stata v.18.

## Data availability

The data that support the findings of this study are available from the corresponding author upon reasonable request; participant-level neurophysiological and clinical data are not publicly available because they contain information that could compromise participant privacy and because the consent obtained did not include public deposition.

## Code availability

Analysis code (Python 3, scipy and statsmodels; and Stata 18) is available from the corresponding author upon reasonable request.

## Acknowledgements

This work was supported by the National Institutes of Health, National Center for Complementary and Integrative Health [grant R01AT009491] and by the Fundação de Amparo à Pesquisa do Estado de São Paulo — FAPESP [grant 2017/12943-8]. The authors thank all participants in both the fibromyalgia and osteoarthritis cohorts for their commitment to the study, and the clinical and research teams at the Spaulding Neuromodulation Center, Harvard Medical School and the Instituto de Medicina Física e Reabilitação, Hospital das Clínicas, Universidade de São Paulo, for their contributions to data collection and management.

## Author contributions

FF: Conceptualization (overall framework, CPRI and INCP constructs, FM study design); Methodology; Formal analysis; Writing original draft; Visualization; Supervision; Funding acquisition; Project administration. MI: Conceptualization (OA study design); Data curation; Writing review & editing; Supervision; Funding acquisition; Project administration. LC: Formal analysis; Data curation; Visualization; Writing — review & editing. KP: Formal analysis; Data curation; Visualization; Writing — review & editing. AG: Data curation; Writing — review & editing. LB: Conceptualization (OA study design); Supervision; Funding acquisition; Project administration; Writing — review & editing. WC: Supervision; Funding acquisition; Project administration; Writing — review & editing. All authors read and approved the final version of the manuscript.

## References

1. Bai Y, Pacheco-Barrios K, Pacheco-Barrios N, Liang G, Fregni F. Neurocircuitry basis of motor cortex-related analgesia as an emerging approach for chronic pain management. Nature Mental Health. 2024;2(5):496–513.

2. Pacheco-Barrios K, Pimenta DC, Pessotto AV, Fregni F. Motor Cortex Inhibition and Facilitation Correlates with Fibromyalgia Compensatory Mechanisms and Pain: A Cross-Sectional Study. Biomedicines. 2023;11(6):1543.

3. Zebhauser PT, Hohn VD, Ploner M. Resting-state electroencephalography and magnetoencephalography as biomarkers of chronic pain: a systematic review. Pain. 2023;164(6):1200–21.

4. Camargo L, Pacheco-Barrios K, Marques LM, Caumo W, Fregni F. Adaptive and Compensatory Neural Signatures in Fibromyalgia: An Analysis of Resting-State and Stimulus-Evoked EEG Oscillations. Biomedicines. 2024;12(7):1428.

5. Viderman D, Kalikanov S, Mukazhan D, Nurmukhamed B. Neurophysiological, Radiological, and Molecular Biomarkers of Pain-Related Conditions: An Umbrella Review. J Clin Med. 2026;15(2):550.

6. Chowdhury NS, Bi C, Furman AJ, Chiang AKI, Skippen P, Si E, et al. Predicting Individual Pain Sensitivity Using a Novel Cortical Biomarker Signature. JAMA Neurol. 2025;82(3):237–46.

7. Shi M, Pacheco LB, Egorova-Brumley N. The effect of depression on the peak alpha frequency as a biomarker of pain sensitivity. Neurobiol Pain. 2025;18:100193.

8. Vockert N, Machts J, Kleineidam L, Nemali A, Incesoy EI, Bernal J, et al. Cognitive reserve against Alzheimer’s pathology is linked to brain activity during memory formation. Nat Commun. 2024;15(1):9815.

9. Le Stanc L, Lunven M, Giavazzi M, Sliwinski A, Youssov K, Bachoud-Lévi AC, et al. Cognitive reserve involves decision making and is associated with left parietal and hippocampal hypertrophy in neurodegeneration. Commun Biol. 2024;7(1):741.

10. McEwen BS, Stellar E. Stress and the individual. Mechanisms leading to disease. Arch Intern Med. 1993;153(18):2093–101.

11. McEwen BS. Allostasis and allostatic load: implications for neuropsychopharmacology. Neuropsychopharmacology. 2000;22(2):108–24.

12. Kahn SE, Hull RL, Utzschneider KM. Mechanisms linking obesity to insulin resistance and type 2 diabetes. Nature. 2006;444(7121):840–6.

13. Borsook D. Allostatic load and pain: instability of a bio-social ecosystem. Pain. 2025;166(11S):S27–S32.

14. Fabris-Moraes W, Lacerda GJM, Pacheco-Barrios K, Fregni F. The Impact of Obesity as a Peripheral Disruptor of Brain Inhibitory Mechanisms in Fibromyalgia: A Cross-Sectional Study. J Clin Med. 2024;13(13):3878.

15. Nijs J, George SZ, Clauw DJ, Fernández-de-las-Peñas C, Kosek E, Ickmans K, et al. Central sensitisation in chronic pain conditions: latest discoveries and their potential for precision medicine. The Lancet Rheumatology. 2021;3(5):e383–e92.

16. Chapman CR, Tuckett RP, Song CW. Pain and stress in a systems perspective: reciprocal neural, endocrine, and immune interactions. J Pain. 2008;9(2):122–45.

17. Ma YX, Lin M, Nan Y, Li NN, Liu B, Wang HR, et al. Obesity-driven low-grade chronic inflammation as a mechanistic bridge to chronic pain: from adipose tissue remodeling to central sensitization. Front Immunol. 2026;17:1889437.

18. Katsuki A, Sumida Y, Gabazza EC, Murashima S, Furuta M, Araki-Sasaki R, et al. Homeostasis model assessment is a reliable indicator of insulin resistance during follow-up of patients with type 2 diabetes. Diabetes Care. 2001;24(2):362–5.

19. Matthews DR, Hosker JP, Rudenski AS, Naylor BA, Treacher DF, Turner RC. Homeostasis model assessment: insulin resistance and beta-cell function from fasting plasma glucose and insulin concentrations in man. Diabetologia. 1985;28(7):412–9.

20. De Ridder D, Adhia D, Vanneste S. The anatomy of pain and suffering in the brain and its clinical implications. Neurosci Biobehav Rev. 2021;130:125–46.

21. Pacheco-Barrios K, Teixeira PEP, Martinez-Magallanes D, Silva Neto M, Pichardo EA, Camargo L, et al. Brain compensatory mechanisms in depression and memory complaints in fibromyalgia: the role of theta oscillatory activity. Pain Med. 2024;25(8):514–22.

22. Barbosa SP, Junqueira YN, Akamatsu MA, Marques LM, Teixeira A, Lobo M, et al. Resting-state electroencephalography delta and theta bands as compensatory oscillations in chronic neuropathic pain: a secondary data analysis. Brain Netw Modul. 2024;3(2):52–60.

23. Schlereth T, Baumgärtner U, Magerl W, Stoeter P, Treede RD. Left-hemisphere dominance in early nociceptive processing in the human parasylvian cortex. Neuroimage. 2003;20(1):441–54.

24. Rinaldi E, van der Heide FCT, Bonora E, Trombetta M, Zusi C, Kroon AA, et al. Lower heart rate variability, an index of worse autonomic function, is associated with worse beta cell response to a glycemic load in vivo—The Maastricht Study. Cardiovasc Diabetol. 2023;22(1):105.

25. Nijs J, Reis F. The Key Role of Lifestyle Factors in Perpetuating Chronic Pain: Towards Precision Pain Medicine. J Clin Med. 2022;11(10):2732.

26. Mickle AM, Tanner JJ, Olowofela B, Wu S, Garvan C, Lai S, et al. Elucidating individual differences in chronic pain and whole person health with allostatic load biomarkers. Brain Behav Immun Health. 2023;33:100682.

27. Picard K, Bisht K, Poggini S, Garofalo S, Golia MT, Basilico B, et al. Microglial-glucocorticoid receptor depletion alters the response of hippocampal microglia and neurons in a chronic unpredictable mild stress paradigm in female mice. Brain Behav Immun. 2021;97:423–39.

28. Lei Y, Wang Q, Wang F, Mu G. Beyond inflammation: a comprehensive microglial regulation model in chronic pain. Mol Biol Rep. 2025;52(1):891.

29. Pribiag H, Stellwagen D. TNF-α downregulates inhibitory neurotransmission through protein phosphatase 1-dependent trafficking of GABA(A) receptors. J Neurosci. 2013;33(40):15879–93.

30. Albrecht DS, Forsberg A, Sandström A, Bergan C, Kadetoff D, Protsenko E, et al. Brain glial activation in fibromyalgia - A multi-site positron emission tomography investigation. Brain Behav Immun. 2019;75:72–83.

31. Simis M, Imamura M, de Melo PS, Marduy A, Pacheco-Barrios K, Teixeira PEP, et al. Increased motor cortex inhibition as a marker of compensation to chronic pain in knee osteoarthritis. Scientific Reports. 2021;11(1):24011.

32. Pacheco-Barrios K, Lima D, Pimenta D, Slawka E, Navarro-Flores A, Parente J, et al. Motor cortex inhibition as a fibromyalgia biomarker: a meta-analysis of transcranial magnetic stimulation studies. Brain Netw Modul. 2022;1(2):88–101.

33. Castelo-Branco L, Uygur Kucukseymen E, Duarte D, El-Hagrassy MM, Bonin Pinto C, Gunduz ME, et al. Optimised transcranial direct current stimulation (tDCS) for fibromyalgia-targeting the endogenous pain control system: a randomised, double-blind, factorial clinical trial protocol. BMJ Open. 2019;9(10):e032710.

34. Simis M, Imamura M, Sampaio de Melo P, Marduy A, Battistella L, Fregni F. Deficit of Inhibition as a Marker of Neuroplasticity (DEFINE Study) in Rehabilitation: A Longitudinal Cohort Study Protocol. Front Neurol. 2021;12:695406.

35. Fregni F, Castelo-Branco L, Cardenas-Rojas A, Daibes M, Silva FM, Pacheco-Barrios K, et al. A randomised, double-blind, sham-controlled, 2×2 factorial trial of aerobic vs. non-aerobic exercise and motor cortex transcranial direct current stimulation in fibromyalgia: effects on clinical outcomes and descending pain modulation. Lancet Reg Health Am. 2026;53:101314.

36. Simis M, Imamura M, Pacheco-Barrios K, Marduy A, de Melo PS, Mendes AJ, et al. EEG theta and beta bands as brain oscillations for different knee osteoarthritis phenotypes according to disease severity. Sci Rep. 2022;12(1):1480.

37. Bennett RM, Friend R, Jones KD, Ward R, Han BK, Ross RL. The Revised Fibromyalgia Impact Questionnaire (FIQR): validation and psychometric properties. Arthritis Res Ther. 2009;11(4):R120.

38. Beck AT, Steer RA, Brown GK. Manual for the Beck Depression Inventory-II. San Antonio, TX: Psychological Corporation; 1996.

39. Zigmond AS, Snaith RP. The hospital anxiety and depression scale. Acta Psychiatr Scand. 1983;67(6):361–70.

40. Bjelland I, Dahl AA, Haug TT, Neckelmann D. The validity of the Hospital Anxiety and Depression Scale. An updated literature review. J Psychosom Res. 2002;52(2):69–77.

41. Buysse DJ, Reynolds CF 3rd, Monk TH, Berman SR, Kupfer DJ. The Pittsburgh Sleep Quality Index: a new instrument for psychiatric practice and research. Psychiatry Res. 1989;28(2):193–213.

42. Johns MW. A new method for measuring daytime sleepiness: the Epworth sleepiness scale. Sleep. 1991;14(6):540–5.

43. World Health Organization. Obesity: preventing and managing the global epidemic. Report of a WHO consultation. World Health Organ Tech Rep Ser. 2000;894:i–xii, 1–253.

44. Khan JS, Hah JM, Mackey SC. Effects of smoking on patients with chronic pain: a propensity-weighted analysis on the Collaborative Health Outcomes Information Registry. Pain. 2019;160(10):2374–9.

45. Carpenter JS, Andrykowski MA. Psychometric evaluation of the Pittsburgh Sleep Quality Index. J Psychosom Res. 1998;45(1):5–13.

46. Murrins Marques L, Pacheco-Barrios K, Camargo L, Houlis M, Vieira J, Castellani A, et al. PIPEMAT-RS: development and validation of a standardized MATLAB pipeline for resting-state EEG preprocessing. J Vis Exp. 2025;(220):e68350.

47. Guidi J, Lucente M, Sonino N, Fava GA. Allostatic Load and Its Impact on Health: A Systematic Review. Psychother Psychosom. 2021;90(1):11–27.

48. McEwen BS. Stress, adaptation, and disease. Allostasis and allostatic load. Ann N Y Acad Sci. 1998;840:33–44.

49. Mhalla A, de Andrade DC, Baudic S, Perrot S, Bouhassira D. Alteration of cortical excitability in patients with fibromyalgia. Pain. 2010;149(3):495–500.

50. Tononi G, Cirelli C. Sleep and the price of plasticity: from synaptic and cellular homeostasis to memory consolidation and integration. Neuron. 2014;81(1):12–34.

51. Nijs J, Loggia ML, Polli A, Moens M, Huysmans E, Goudman L, et al. Sleep disturbances and severe stress as glial activators: key targets for treating central sensitization in chronic pain patients? Expert Opin Ther Targets. 2017;21(8):817–26.

52. Vidor LP, Torres IL, Medeiros LF, Dussán-Sarria JA, Dall’agnol L, Deitos A, et al. Association of anxiety with intracortical inhibition and descending pain modulation in chronic myofascial pain syndrome. BMC Neurosci. 2014;15:42.

53. Thompson JM, Neugebauer V. Cortico-limbic pain mechanisms. Neurosci Lett. 2019;702:15–23.

54. Snow NJ, Kirkland MC, Downer MB, Murphy HM, Ploughman M. Transcranial magnetic stimulation maps the neurophysiology of chronic noncancer pain: a scoping review. Medicine (Baltimore). 2022;101(46):e31774.

55. González-Roldán AM, Cifre I, Sitges C, Montoya P. Altered dynamic of EEG oscillations in fibromyalgia patients at rest. Pain Med. 2016;17(6):1058–68.

56. Ji G, Neugebauer V. Hemispheric lateralization of pain processing by amygdala neurons. J Neurophysiol. 2009;102(4):2253–64.

57. Bollen K, Lennox R. Conventional wisdom on measurement: a structural equation perspective. Psychol Bull. 1991;110(2):305–14.

58. Long JS, Ervin LH. Using heteroscedasticity consistent standard errors in the linear regression model. Am Stat. 2000;54(3):217–24.

