## Supplementary Note for "Motor cortex pain compensation is modifiable: allostatic load degrades its analgesic efficiency and rehabilitation restores it"

**Supplementary Note 1. Preliminary clinical calibration and implementation feasibility of the CPRI**

All values establish the order of magnitude of the index’s performance and provide anchors for prospective calibration; they are not clinical decision rules. Because CPRI is normalised to a cohort mean of 1.0 using cohort-specific parameters, every absolute value denotes a position within these cohorts, not an absolute physiological quantity. The standardisation, shift and normalisation constants were estimated from the baseline distribution and applied unchanged to post-treatment data, so post-treatment values are not re-centred and change scores are interpretable on the baseline scale.

**Supplementary Table 1. Distributional anchors for the CPRI, by cohort.**

|  | **Fibromyalgia** | **Osteoarthritis** |
| --- | --- | --- |
| Participants with complete pre- and post-treatment CPRI | 73 | 54 |
| Participants with classifiable clinical response | 73 | 54 |
| Response criterion* | FIQR improvement ≥14 points | SF-36 PF improvement ≥10 points |
| Responders / non-responders | 23 / 50 | 25 / 29 |
| Cohort mean (SD) | 1.00 (0.47) | 1.00 (0.75) |
| Skewness | −0.03 | 1.07 |
| INCP− mean (SD) | 0.85 (0.45) | 0.76 (0.62) |
| INCP+ mean (SD) | 1.17 (0.44) | 1.20 (0.80) |
| INCP+ vs INCP−: *t*; *p*; *d* | 3.06; 0.003; 0.72 | 2.53; 0.014; 0.62 |

CPRI, Cortical Pain-Compensatory Resistance Index; FIQR, Fibromyalgia Impact Questionnaire–Revised; INCP, Impaired Neurocompensatory Capacity Phenotype; SF-36 PF, 36-Item Short Form Health Survey Physical Functioning subscale. Distribution statistics use all participants with complete pre- and post-treatment data; response-based analyses are restricted to participants with a classifiable response status. Group comparisons are two-sided Welch t-tests.

*Response criteria were pre-specified. The ≥14-point FIQR threshold is a conservative, distribution-based criterion and is not an established FIQR minimal clinically important difference: the published 14% relative-change value was derived for the original FIQ rather than the FIQR, and no anchor-based minimal clinically important difference has been established for the FIQR (reported minimal detectable change values range from approximately 15 to 22 points). The ≥10-point SF-36 PF threshold lies within the 8–10-point range commonly reported for that subscale but has not been derived in osteoarthritis.

Pooled fixed-effects estimate for CPRI elevation by phenotype: d = 0.67 (95% CI 0.33–1.02), p = 0.00014.

**Supplementary Table 2. Change in CPRI by clinical response, and provisional minimum important change.**

|  | **Fibromyalgia** | **Osteoarthritis** |
| --- | --- | --- |
| ΔCPRI, responders, mean (SD) | −0.85 (0.43) | −0.64 (0.69) |
| ΔCPRI, non-responders, mean (SD) | +0.05 (0.48) | +0.17 (0.70) |
| Welch *t*; *p*; *d*† | 7.91; <0.0001; −1.91 | 4.30; <0.0001; −1.17 |
| ½ baseline SD | 0.235 | 0.375 |
| **Provisional minimum important change** | **0.25 units** | **0.25 units** |

†Negative values of *d* denote a greater reduction in CPRI among responders than non-responders. Because clinical response is defined by change in the symptom measure that also enters the CPRI, this contrast is presented as a descriptive consistency check rather than as independent evidence of neurophysiological change; the corresponding non-circular result is the difference in ΔPCS by response status reported in the main text.

**Preliminary response threshold. Receiver-operating-characteristic analysis was performed in the fibromyalgia cohort (n = 73). Baseline CPRI discriminated clinical response with an area under the curve of 0.70 (approximate 95% CI 0.56–0.84), and the Youden-optimal threshold of CPRI = 1.10 gave a sensitivity of 0.70 and a specificity of 0.68; the response rate was 46% (17/37) above a CPRI of 1.0 and 17% (6/36) at or below it. The threshold was derived and evaluated in the same sample without internal validation or optimism correction, and is offered as a calibration anchor rather than a decision rule.**

**Calibration at a new centre. A centre may either apply the standardisation and shift constants published here — yielding values comparable to Supplementary Table 1, but requiring close protocol matching — or estimate its own parameters locally, yielding an internally valid but not externally comparable index. The latter is the route HOMA-IR takes, since insulin assays require laboratory-specific calibration. One departure from the metabolic analogy should be stated explicitly: fasting glucose and insulin are measured on absolute, transferable scales, whereas both CPRI components are standardised and shifted within cohort. An individual’s CPRI therefore depends on the distribution of the sample in which it is computed, and the values reported for the two cohorts here are not directly comparable to one another.**

**Minimum important change. The value of 0.25 units is distribution-based, following the half-standard-deviation heuristic,1 and is derived from the fibromyalgia baseline distribution (half the fibromyalgia SD is 0.235; half the osteoarthritis SD is 0.375; half the pooled SD is 0.31). A single provisional value is proposed across cohorts for simplicity; a cohort-specific value for osteoarthritis would be 0.375. It is not anchor-based and has not been compared with the minimum detectable change, since test–retest reliability of the index is not yet established; anchor-based derivation and a test–retest study are the appropriate next steps.**

**Implementation feasibility. The two inputs face unequal constraints. The theta-asymmetry term needs only two central electrodes, and dry-electrode systems agree well with gel-based research systems in the theta band specifically (r ≈ 0.80),2 so a low-channel implementation is plausible — although most consumer headsets omit C3 and C4 altogether, and a difference between homologous electrodes is the metric most sensitive to inter-electrode impedance mismatch, making device-specific validation a prerequisite. The SICI term is the harder constraint: portable stimulators now exist,3 but they deliver therapeutic repetitive stimulation, whereas short-interval intracortical inhibition requires paired-pulse hardware, electromyography and a trained operator. A simplified index would therefore depend either on the EEG term alone retaining prognostic performance, or on substituting an EEG-derived index of cortical excitation–inhibition balance4 for paired-pulse SICI — both testable in these data, neither yet established.**

**References**

*References in this Supplementary Information are numbered separately from those in the main text.*

1. Norman GR, Sloan JA, Wyrwich KW. Interpretation of changes in health-related quality of life: the remarkable universality of half a standard deviation. *Med Care* 2003; 41: 582–92.

2. Kleeva D, Ninenko I, Lebedev MA. Resting-state EEG recorded with gel-based vs. consumer dry electrodes: spectral characteristics and across-device correlations. *Front Neurosci* 2024; 18: 1326139.

3. Qi Z, Liu H, Jin F, et al. A portable transcranial magnetic stimulation device. *Nat Commun* 2025; 16: 2731.

4. Gao R, Peterson EJ, Voytek B. Inferring synaptic excitation/inhibition balance from field potentials. *NeuroImage* 2017; 158: 70–8.
